# Swarm-GestaltMatcher: distributed Gestalt learning through Swarm Learning to enhance facial phenotyping for rare genetic syndromes

**DOI:** 10.64898/2026.01.02.26343337

**Authors:** Arnab Bandyopadhyay, Quentin Hennocq, Olivier Lienhard, Tzung-Chien Hsieh, Ahmed Zaiter, Luan Breton, Stefanie Warnat-Herresthal, Hartmut Schultze, Anna Aschenbrenner, Valérie Cormier-Daire, Roman Hossein Khonsari, Joachim L. Schultze, Peter Krawitz

## Abstract

Deep learning–based facial phenotyping represents a major paradigm shift in the diagnosis of rare and ultra-rare genetic disorders. By capturing disease-specific craniofacial “gestalts” that are often subtle, overlapping, but overlooked in routine clinical practice, these technologies surpass the traditional limits of dysmorphology assessment. Despite this, data scarcity and stringent privacy policies constraint centralized model training and its clinical translation. Swarm learning, a decentralized paradigm that combines edge-based training with blockchain-mediated parameter synchronization, offers a potential solution by enabling collaborative model development without sharing raw patient data. However, it remains uncertain whether a decentralized model can achieve performance comparable to the centralized model, particularly in the context of rare disease diagnosis. Using two complementary datasets, GMDB, and AIDY, we evaluate the feasibility, performance, and clinical relevance of swarm learning against centralized and institution-specific local models. Model performance is assessed across in-distribution, cross-institutional, and ultra-rare disorder scenarios (not part of model training), with additional analyses of calibration and epistemic uncertainty. Our results show that swarm-trained models consistently match the accuracy of centralized training and outperformed local models. Importantly, swarm learning preserves sensitivity to low-prevalence and ultra-rare syndromes despite extreme data scarcity across sites, while exhibiting more conservative and reliable uncertainty calibration. Although swarm learning has previously been applied to well-characterized diseases, this study represents the first application of a swarm model in real-world settings involving a large and diverse set of disease classifications. Taken together, swarm learning emerges as a scalable, equitable, and trustworthy framework that shortens the diagnostic odyssey and advances precision medicine for rare disease diagnosis in routine clinical practice.

## Introduction

Rare genetic disorders (RGDs) impact an estimated 3.5–5.9% of the global population (1), with over 7000 distinct conditions catalogued in the Online Mendelian Inheritance in Man (OMIM) database and nearly 5000 genes are associated with mutations that result in clinically observable phenotype (2). Diagnostic failure is common due to limited routine screening for non-coding or structural variants (3). Emerging data suggest that a disease phenotype arise from complex pathobiological interactions within the interactome (4,5), where perturbation of functionally related genes can result to overlapping clinical phenotype (6). Identifying gene-disease relationships in ultra-rare disorders is even more challenging due to the limited case reports available. Many RGDs exhibit recurrent craniofacial patterns or a recognizable “gestalt” that can provide critical diagnostic cues. Nevertheless, craniofacial phenotyping remains complex, often subjective and examiner-dependent(7). The process of differential diagnosis is further confounded by the vast number of rare genetic syndromes (8). Consequently, diagnosis is challenging and time-consuming, often the waiting period is over five years. This is often referred to as the “diagnostic odyssey” (9).

Recent advances in AI-based facial phenotyping technologies have significantly enhanced diagnostic accuracy in this domain (9–11). Several next-generation phenotyping (NGP) tools have been developed (11–20) that employ computer-aided facial analysis using standard two-dimensional patient photographs or three-dimensional imaging, providing a powerful tool for improving clinical decision-making. Among the NGP tools, a land mark work is DeepGestalt (19) that supports ∼216 genetic disorders. Recently, GestaltMatcher was proposed as an extension of DeepGestalt (21,22). This approach converts each frontal facial image into a high-dimensional embedding that quantitatively represents dysmorphic facial anatomy. These embeddings represent a continuous anatomical–phenotypic feature space, called ’Clinical Face Phenotype Space’ (CFPS) (21). In CFPS, patients can be matched and clustered based on shared craniofacial morphology, independent of predefined diagnostic labels.

Even though GestaltMatcher enables faster, more consistent recognition of known syndromes, it is trained in a centralized fashion, limited in number of patients, images and disorders. Centralized method necessitates aggregation of large-scale, multi-institutional datasets at a single site. This data consolidation is severely constrained by patient privacy regulations, institutional governance policies, cross-border data transfer restrictions, GDPR and ethical considerations. In addition, centralized method implicitly overrepresents resource enrich centres and marginalize low-resource or unique populations. In such scenario, model overfits dominant disease and suffers from poor performance for low-prevalence, ultra-rare disorders. This limits anatomical generalizability. In addition, important drawback of centralized training is the model’s ability to continuously adapt to evolving data distributions, newly emerging syndromes, and rare phenotypic variants encountered in routine clinical practice. Model updates require repeated large-scale data transfers and retraining cycles, which are computationally expensive and poorly scalable.

To tackle these limitations and to advance the current state of GestaltMatcher algorithms, we opt out for a decentralized swarm learning method. Recently swarm learning has emerged as an alternative to the centralized learning and have used in various disease conditions (23–27). It enables multiple institutions to collaboratively train a deep learning model without exchanging raw patient data. It synchronizes model parameters through a blockchain-enabled, peer-to-peer network with dynamic leader election. This architecture ensures data locality, cryptographic security, fault tolerance, and institutional data sovereignty, while enabling continuous model improvement across geographically and administratively independent distributed sites. Importantly, this method increases the training cohort size for ultra-rare syndromes that are otherwise underrepresented within any single center.

In this work, we introduced two key methodological innovations. First, we shifted from the conventional centralized training method to decentralized Swarm Learning. Second, we trained the model jointly on two independent and complementary datasets: GMDB (GestaltMatcher Data Base) and AIDY (Artificial Intelligence for Dysmorphology). Our results indicated that the swarm model exhibited more conservative predictions, while the central model tended to be more overconfident in some cases. We further demonstrated that both, Swarm and Central models achieved similar accuracies when trained on both datasets.

## Results and Discussion

### Two distinct facial imaging databases increase number and diversity of rare disease syndromes

In this work, we considered two datasets, Gestalt Matcher Data Base (GMDB, v1.1.0) and AIDY dataset. Both datasets employ a version control system for tracking changes and managing files. GestaltMatcher Database collects medical images of individuals with clinically or molecularly confirmed diagnoses from publications. AIDY data contains images from photographic archives of the Maxillofacial Surgery, Plastic Surgery, and Medical Genetics departments at Necker–Enfants Malades Hospital (AP-HP). A comprehensive description of datasets, preprocessing, quality-control procedures, and additional methodological details is provided in the Methods section. For consistency and comparability, we adopted the same definitions of Frequent and Rare syndromes and the same data-splitting strategy as used in GestaltMatcher (see ref 22, 23). Specifically, a syndrome was categorized as Frequent if it contained more than six images. Syndromes with six or fewer images were categorized as Rare. For both categories, we applied a 90:10 stratified split for training and testing. Table 1 summarizes the overall composition of the AIDY and GMDB datasets in terms of the number of images, patients, and syndromes. Overall, the AIDY dataset contains a larger total number of images in both the Frequent and Rare subsets. Conversely, GMDB exhibits greater diversity in the number of distinct syndromes and patients. Consequently, AIDY includes more images per patient, whereas GMDB provides fewer images per patient but a broader representation of syndromic diversity.

**Table 1.** Dataset comparison in number of images, patients and disorders.

| Table 1. Dataset comparison in number of images, patients and disorders |  |  |  |  |  |  |
| --- | --- | --- | --- | --- | --- | --- |
|  | GMDB |  |  | AIDY |  |  |
| Dataset | # images | # patients | # disorders | # images | # patients | # disorders |
| Frequent | 9,693 | 7,314 | 277 | 26,237 | 2,967 | 109 |
| Rare | 1,287 | 1,032 | 299 | 4,332 | 589 | 242 |
| Total | 10,980 | 8,346 | 576 | 30,569 | 3,540 | 351 |

Figure S1 displays the disorder distribution for AIDY (A), GMDB (B) and together (C). The x-axis represents the number of patients per disorder, while the y-axis (logarithmic scale) indicates the number of disorders with the corresponding number of patients in the X-axis. The black bars correspond to the Rare subsets (AIDY-Rare or GMDB-Rare), defined by disorders with more than one but fewer than seven patients. Both datasets exhibit a long-tail distribution, with substantially more rare syndromes, 299 in GMDB and 242 in AIDY. Together it comprises of total 385 Rare disorders. Only a small number of disorders have large patient counts. In GMDB-Rare, 12.4% of patients account for 51.9% of disorders and 11.7% of images. In AIDY-Rare, 16.6% of patients account for 68.9% of disorders and 14.1% of images.

To investigate whether the AIDY data contains syndrome that are underrepresented in GMDB and vice versa, we harmonized syndrome names across datasets (see methods). We identified 40 common syndromes classified as Rare in both datasets. Additionally, 88 syndromes are Rare in AIDY dataset but Frequent in GMDB and 15 syndromes are Rare in GMDB but Frequent in AIDY dataset. Interestingly, in Frequent category, we found 34 unique syndromes in AIDY and 129 in GMDB. This underscores the complementary nature of the two datasets. Details of disorders common or uncommon across both datasets, and those rare in one but frequent in the other, are included in the supplementary Table S1. Disease identifiers are provided as OMIM and ORPHA codes, and the corresponding disease names can be retrieved from their respective databases.

In Figure S1D we illustrate ethnic diversity for both datasets. In both dataset, European ancestry population represent a substantial proportion, >30% and >50% in AIDY and GMDB, respectively. Representation of individuals of African ancestry is limited in both datasets (∼3% in AIDY and ∼6% in GMDB). Sex distribution is approximately balanced in both datasets, at the level of patients and images. However, the datasets differ notably in age distribution. Nearly half of GMDB images correspond to infants, whereas only approximately 10% of AIDY images fall into this age group. Conversely, AIDY contains a higher proportion (∼35%) of images from individuals aged 10 years or older, compared to approximately 20% in GMDB.

Overall, our exploratory analyses reveal that AIDY and GMDB complement each other in both phenotypic and demographic dimensions. While AIDY provides larger image counts per patient, more mature faces, GMDB offers broader syndromic and patient diversity, infant (developmental) faces. The datasets also include substantial numbers of mutually unique syndromes. Therefore, joint model training leveraging both datasets is expected to combine their respective strengths, improve generalizability, and enhance coverage of rare and heterogeneous disorders.

### Swarm model achieves comparable performance to Centralized model in rare disease classification

Because the GMDB dataset exhibits greater diversity in both the number of patients and the range of syndromes, we selected it to evaluate the feasibility of decentralized Swarm Learning. To ensure consistency and enable a fair comparison between the swarm-trained and centrally trained models, we adopted the same evaluation protocol used in GestaltMatcher (23). Specifically, the identical backbone architecture, iResNet-50 and iResNet-100 pretrained on the Glint360K dataset was fine-tuned on GMDB. In addition, model ensemble and test-time augmentation (TTA) were enforced as in GestaltMatcher. For inference, averaged cosine distance of all models in the ensemble and TTAs per model between the gallery images to the test image was used to rank the disorders. With three models and two augmentations per model, this yielded 12 cosine distance measurements per query image, which were then used to rank candidate disorders.

For the first experiment, the GMDB Frequent and Rare dataset was split into Gallery and Test set in 90:10 ratio. For the swarm experiment, Frequent Gallery set was splited in a stratified manner across two swarm nodes. One node therefore contains half of the Gallery data and only this amount of data is visible to the Swarm model during training. The other node contains other half of the Gallery data and similar settings applied. During fine-tuning, both the centralized and decentralized models were trained using weighted cross-entropy loss. All models were trained for 50 epochs. In the swarm setup, the synchronization interval was configured such that parameter aggregation occurred approximately every five epochs, resulting in around ten synchronization events during training. Evaluation was done using the completely held-out Test set. Although different synchronization intervals were explored during training, model performance remained largely unchanged. Full details are provided in the Methods section.

A comparison of diagnostic performance between the centralized model and the Swarm model is presented in Figure S2. Dotted line with square and circle represent corresponding error in prediction. Dotted triangle represents gain in accuracy between Top-K. For the Frequent testing set, we observed marginal performance gain in Centralized training method in Top 5 (∼5%), Top 10 (∼2%) and Top 30 (∼2%) category. Top 1 performance remains same for Swarm and Central model. When the model is supplied Frequent and Rare Gallery images, we observed a similar trend in performance drop in Central and Swarm model compared to when only Frequent Gallery set was supplied. However, marginal performance gain (∼2-5%) is observed in Centralized method similar to the previous case. Comparing performance for Rare set is interesting as this set is was not part of the model training and can infer about the generalizibility of the model. We expected that the generalizibility of Swarm model would be significantly different compared to the Central model as these set was not part of the training and in each node, model only seen half of the Frequent Gallery data. We observed that the model generalizes very similar to the central model and as in the previous case ∼2-4% performance gain is observed in Central model. As reported before, we observed performance drop for central model for rare test set when Frequent and rare gallery was supplied. This is also visible for Swarm model. However, in this case the performance gain between these two models remains within the range of previous case.

Using a stratified data split, we observed only a modest performance difference of approximately 2–5% between Central and Swarm model in both Frequent and Rare diseases. Additionally, Swarm model shows comparable generalization on the Rare set and yielded similar performance. To further stress-test model robustness under extreme data heterogeneity, we evaluated a non-stratified sex-based split. In this set-up, one node contained only male samples and the other only female samples. In Figure S3 we compare model performances for Frequent and Rare disorders. For Frequent disorders, the central model showed an approximately 5-10% performance advantage in the Top-1 and Top-5 metrics. However, for Rare disorders, performance of both models remains comparable. Further, we evaluated model performance on held-out male and female test sets. Under this setting, the central model showed an approximately 11% performance advantage on male samples in the Top-1 and Top-5 metrics (Figure S4). However, this gap narrowed to around 2% for the Top-10 and Top-30 metrics. For female samples, the central model demonstrated a performance gain of roughly 7% in the Top-1, Top-5, and Top-10 categories, which similarly decreased to approximately 3% at the Top-30 level. A further ethnicity-based split across two nodes yielded comparable trends, with swarm-trained models maintaining performance close to centralized training. Collectively, these results indicate that while centralized models may retain a slight advantage under highly skewed demographic splits at strict ranking thresholds, swarm model remains robust and competitive.

### Swarm model preserves Centralized-level performance in real-world, cross-institutional training while outperforming local models

Even though there is a performance gap between central and swarm model, we demonstrated that Swarm model performs and generalizes similarly as the central model. Now we would like to train Swarm model in real world setting where GMDB data and AIDY data are hosted in two separate nodes. We have used the same definition of Frequent and Rare diseases as in GMDB and then further split into Gallery and test images in 90:10 ratio. We found in exploratory analysis that some diseases present in GMDB but not in AIDY and vice versa. Therefore, for fair evaluation of GMDB and AIDY model on AIDY and GMDB test data, we have used only diseases that are common in both datasets.

To systematically assess the relative performance of decentralized training versus local and centralized method, we evaluated four distinct models across four ranking-based metrics (Top-1, Top-5, Top-10, and Top-30 accuracy) for both frequent and rare disorder classification. The first two models are local models, namely the GMDB model and the AIDY model, each trained exclusively on their respective institutional datasets. The third model is a centralized model, trained by gathering GMDB and AIDY data into a single repository. The fourth model is a decentralized swarm model, in which GMDB and AIDY data remain strictly local and fully isolated like in a real-world, privacy-preserved scenario. In this case model parameters are periodically synchronized at predefined intervals using a decentralized coordination protocol. In addition to in-distribution evaluation (GMDB->GMDB or AIDY->AIDY), we checked cross-data generalization to assess robustness under domain shift and data heterogeneity. For example, the GMDB model was evaluated on held-out AIDY test data (GMDB->AIDY), and conversely, the AIDY model was evaluated on held-out GMDB data (AIDY->GMDB). Furthermore, both the centralized and swarm models were independently evaluated on the GMDB (centralized->GMDB, swarm->GMDB) and AIDY held-out test sets (centralized->AIDY, swarm->AIDY). This evaluation framework enables a rigorous comparison of local, centralized, and decentralized models in terms of diagnostic accuracy, rare disease sensitivity, and cross-institutional generalizability. Figure 2 and Table 2 summarize all evaluation metrices used in the study.

**Figure 1.**
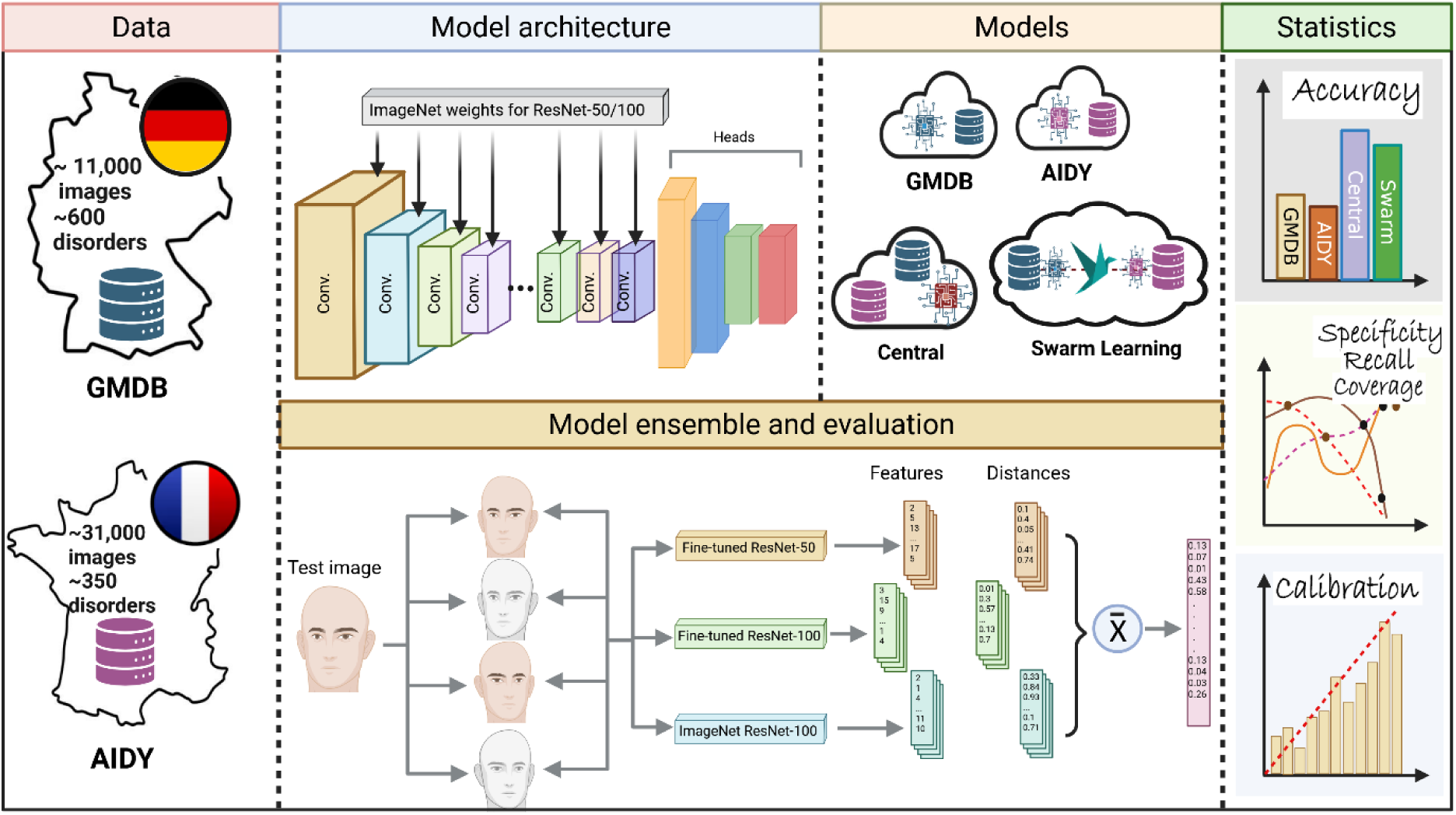
Schematic overview of the methodology used in this study. Data from GMDB and AIDY were used to fine-tune ImageNet face recognition models of various depths. Data augmentation and model ensemble methods were applied to evaluate local models (GMDB and AIDY), as well as Central and Swarm Learning models, and various performance metrices were computed.

**Figure 2.**
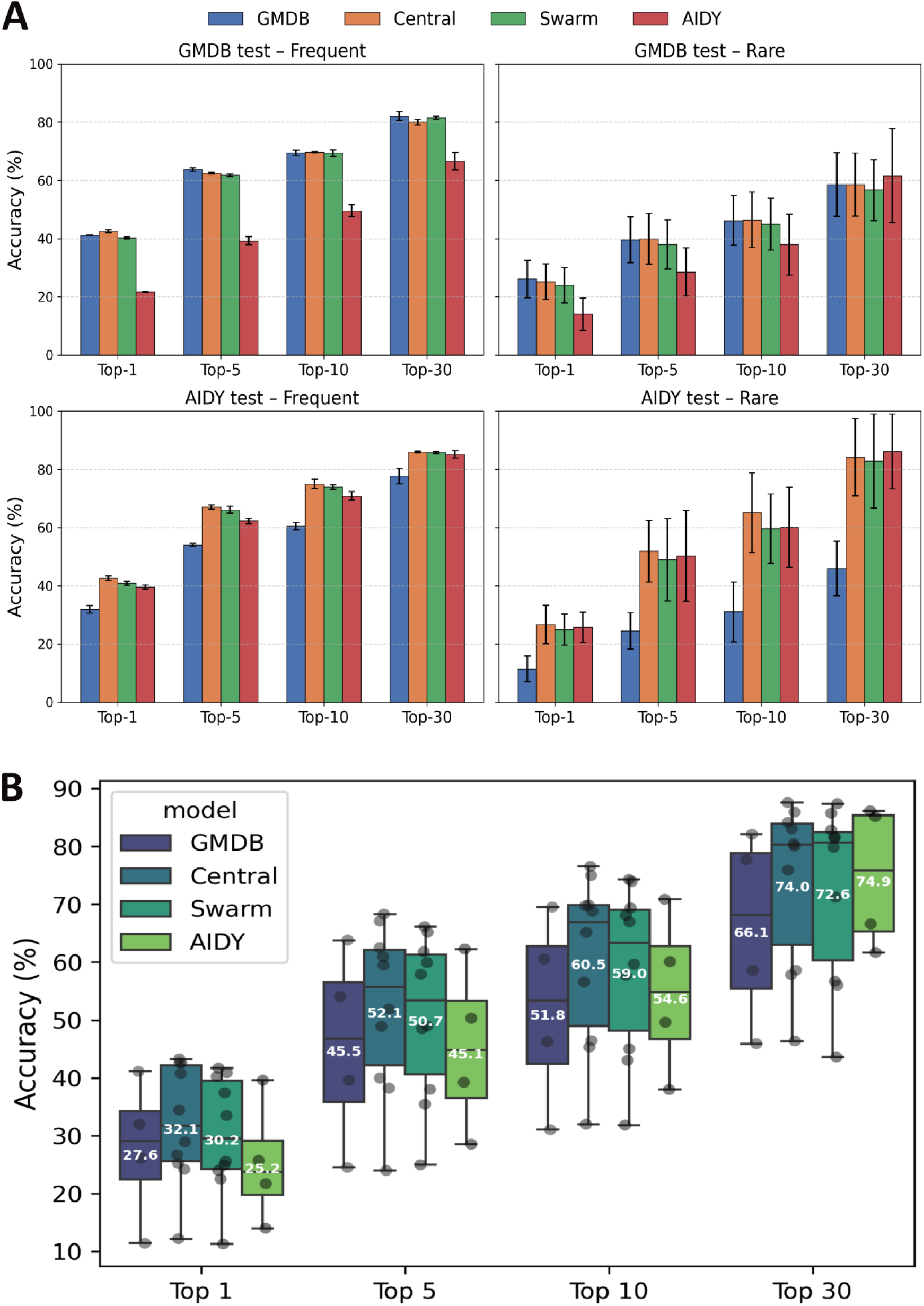
A) GMDB, AIDY, Central and Swarm model comparison on local and cross-institutional data and Top 1, Top 5, Top 10, and Top 30 accuracies are plotted for Frequent (left panels) and Rare (right panels) disorders. B) The average performance of all models across all datasets and disease categories (Frequent and Rare) are shown for Top K evaluation. Mean values for each model are represented in white.

**Table 2.** Performance of different model on GMDB Frequent and Rare disorders.

| Model -> data | Category<br>(Per syndrome<br>mean) | Top 1 | Top 5 | Top 10 | Top 30 |
| --- | --- | --- | --- | --- | --- |
| GMDB -> GMDB | Frequent/Frequent | 41.23 | 64.43 | 70.46 | 83.66 |
| AIDY -> GMDB | Frequent/Frequent | 21.83 | 40.57 | 51.70 | 69.57 |
| Central -> GMDB | Frequent/Frequent | 43.04 | 62.77 | 70.02 | 81.00 |
| Swarm -> GMDB | Frequent/Frequent | 40.52 | 62.25 | 70.51 | 82.13 |
| GMDB -> GMDB | Frequent/Overall | 40.97 | 63.17 | 68.59 | 80.58 |
| AIDY -> GMDB | Frequent/Overall | 21.56 | 37.84 | 47.49 | 63.61 |
| Central -> GMDB | Frequent/Overall | 42.05 | 62.2 | 69.45 | 79.11 |
| Swarm -> GMDB | Frequent/Overall | 39.9 | 61.32 | 68.22 | 80.99 |
| GMDB -> GMDB | Rare/Rare | 32.5 | 47.42 | 54.79 | 69.54 |
| AIDY -> GMDB | Rare/Rare | 19.56 | 36.79 | 48.41 | 77.79 |
| Central -> GMDB | Rare/Rare | 31.30 | 48.65 | 55.94 | 69.43 |
| Swarm -> GMDB | Rare/Rare | 30.02 | 46.47 | 53.91 | 67.18 |
| GMDB -> GMDB | Rare/Overall | 19.71 | 31.74 | 37.71 | 47.60 |
| AIDY -> GMDB | Rare /Overall | 8.38 | 20.26 | 27.48 | 45.52 |
| Central -> GMDB | Rare /Overall | 19.13 | 31.23 | 36.92 | 47.74 |
| Swarm -> GMDB | Rare /Overall | 17.93 | 29.52 | 36.11 | 46.18 |

**Table 3.** Performance of different model on AIDY Frequent and Rare disorders

| Model -> data | Category<br>(Per syndrome<br>mean) | Top 1 | Top 5 | Top 10 | Top 30 |
| --- | --- | --- | --- | --- | --- |
| GMDB -> AIDY | Frequent/Frequent | 33.22 | 54.52 | 61.76 | 80.43 |
| AIDY -> AIDY | Frequent/Frequent | 40.22 | 63.24 | 72.33 | 86.41 |
| Central -> AIDY | Frequent/Frequent | 43.38 | 67.82 | 76.58 | 86.23 |
| Swarm -> AIDY | Frequent/Frequent | 41.56 | 67.28 | 74.79 | 86.15 |
| GMDB -> AIDY | Frequent/Overall | 30.61 | 53.58 | 59.31 | 75.05 |
| AIDY -> AIDY | Frequent/Overall | 38.93 | 61.28 | 69.38 | 83.90 |
| Central -> AIDY | Frequent/Overall | 41.85 | 66.35 | 73.39 | 85.67 |
| Swarm -> AIDY | Frequent/Overall | 40.18 | 65.00 | 73.10 | 85.37 |
| GMDB -> AIDY | Rare/Rare | 15.81 | 30.67 | 41.28 | 55.25 |
| AIDY -> AIDY | Rare/Rare | 30.86 | 65.91 | 73.83 | 99.07 |
| Central -> AIDY | Rare/Rare | 33.33 | 62.47 | 78.85 | 97.47 |
| Swarm -> AIDY | Rare/Rare | 30.24 | 63.19 | 71.56 | 99.07 |
| GMDB -> AIDY | Rare/Overall | 6.97 | 18.30 | 20.74 | 36.54 |
| AIDY -> AIDY | Rare /Overall | 20.55 | 34.65 | 46.34 | 73.26 |
| Central -> AIDY | Rare /Overall | 20.07 | 41.24 | 51.40 | 70.89 |
| Swarm -> AIDY | Rare /Overall | 19.56 | 34.70 | 47.72 | 66.62 |

In Figure 2A, the upper and lower panels show model performances on the GMDB and AIDY test datasets, respectively, for frequent diseases (left) and rare diseases (right). The swarm-trained model demonstrates performance that is comparable to the centralized model across Top-5, Top-10, and Top-30 accuracy for frequent disorder classification. Notably, the swarm model consistently outperforms the corresponding local models on their own institutional test sets, for example, in the AIDY→AIDY evaluation. In cross-institutional testing, the GMDB model evaluated on AIDY data (Figure 2A, lower panel) achieves reasonable generalization but does not surpass the swarm model. A similar trend is observed when the AIDY model is evaluated on GMDB data. For rare disorder classification, the swarm model achieves performance comparable to the local models. Specifically, for AIDY rare disorders, the swarm model performs similarly to both the GMDB and AIDY local models. Likewise, for GMDB rare disorders, comparable performance is observed between the GMDB local model and the swarm model. This indicates that swarm training does not compromise sensitivity to low-prevalence conditions.

Overall, for frequent disorders, the difference between centralized and swarm training is below 1% in most metrics. In this category, swarm model demonstrates better performance compared to the local models. For rare disorders, the difference between central and swarm model remains below 3% for all metrics. Collectively, these results demonstrate that swarm learning enables effective integration of distributed data without sacrificing diagnostic performance, for both frequent and rare disease categories, while preserving institutional data sovereignty and privacy.

In Figure 2B, we report the average performance of all models across cross-institutional datasets and disease categories (Frequent and Rare) for Top-K evaluation. The White values represent mean values. The Swarm model outperformed the local models, with performance gains ranging from 3-7%. The performance difference between the Swarm model and centralized model was around 1.5%.

### Swarm model is conservative in nature

We additionally evaluated the probabilistic calibration of both the centrally trained model and the Swarm model using the reliability diagram and expected calibration error (ECE). ECE provides a quantitative measure of how well a model’s predicted confidence aligns with its observed empirical accuracy. Proper calibration is essential for clinical decision-support systems, as it directly influences how much trust a clinician can place in the model’s confidence scores when interpreting diagnostic suggestions. For this comparison, we evaluated the ResNet-100 model trained in Centralized manner and Swarm learning model, as the ResNet-100 architecture demonstrated superior performance compared to the ResNet-50 model. For each input image, the model produced a probability distribution across disease classes, and the maximum softmax probability was selected as the prediction confidence. Prediction accuracy was determined by comparing the predicted labels with the ground-truth labels.

Confidence scores are grouped into 30 equally spaced bins spanning the interval [0, 1]. For each bin 𝐵_𝑚_, the average confidence and empirical accuracy were calculated as

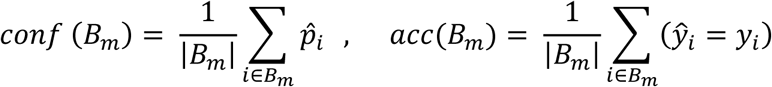

Where *p̂_i_* denotes the predicted confidence, 𝑦^_𝑖_ the predicted class, and 𝑦_𝑖_ the true class label for sample 𝑖. Reliability diagrams were constructed by plotting the empirical accuracy against the predicted confidence for each bin. The black dashed diagonal line represents perfect calibration, where predicted confidence and actual accuracy are in exact agreement. Blue bars represent the empirical accuracy within each confidence bin, while the shaded gap between confidence and accuracy illustrated calibration error. Red scatter points indicated the mean confidence and accuracy for each bin. To quantify statistical uncertainty, Wilson score confidence intervals were computed for the bin-wise accuracies and displayed as error bars. This approach provides more robust interval estimates, particularly for bins with smaller sample sizes. Expected Calibration Error (ECE) was computed as the weighted average of the absolute difference between bin accuracy and confidence.

In Figure 3, predicted confidence is plotted on the x-axis and the corresponding observed accuracy on the y-axis for the centralized model (left panel) and the Swarm model (right panel). The centralized model demonstrates good adherence to this ideal relationship in the low- and mid-confidence ranges. However, at higher confidence regions, the empirical accuracy consistently falls below the diagonal, indicating systematic overconfidence. This model reports high certainty even when its predictions are not proportionally reliable. Conversely, the swarm-trained model exhibits the opposite pattern. Across confidence bins, its empirical accuracy remains above the perfect-calibration line, reflecting a conservative calibration profile. In this scenario, the model’s true accuracy exceeds its stated confidence. Such conservative behaviour is advantageous in clinical settings, as it mitigates the likelihood of a highly confident but incorrect classification, thereby reducing potential diagnostic risk. To further investigate the calibration gap observed in the Central model, we categorised diseases into two groups: high-representation (>50 images) and low-representation (<50 images). For high-representation diseases, the Central model performs close to the ideal calibration line. However, for low-representation diseases, a substantial deviation from the ideal line is evident, especially in the high-confidence region. It is likely that the Central model learns strong confidence patterns from frequent classes but lacks enough diverse examples to properly calibrate uncertainty for rare conditions, leading to overconfident errors. As a result, for underrepresented diseases, the Central model remains highly confident despite reduced predictive accuracy. By comparison, the Swarm model maintains consistent behaviour across both subgroups. Its empirical accuracy remains above the perfect-calibration line for both high-and low-representation diseases, further supporting its conservative calibration profile. In contrast to the Central model, the Swarm model benefits from knowledge shared across multiple institutions, exposing it to broader disease diversity and variation. This richer training signal improves generalisation and helps the model remain cautious when evidence is limited, resulting in a more conservative and clinically safer calibration profile.

**Figure 3.**
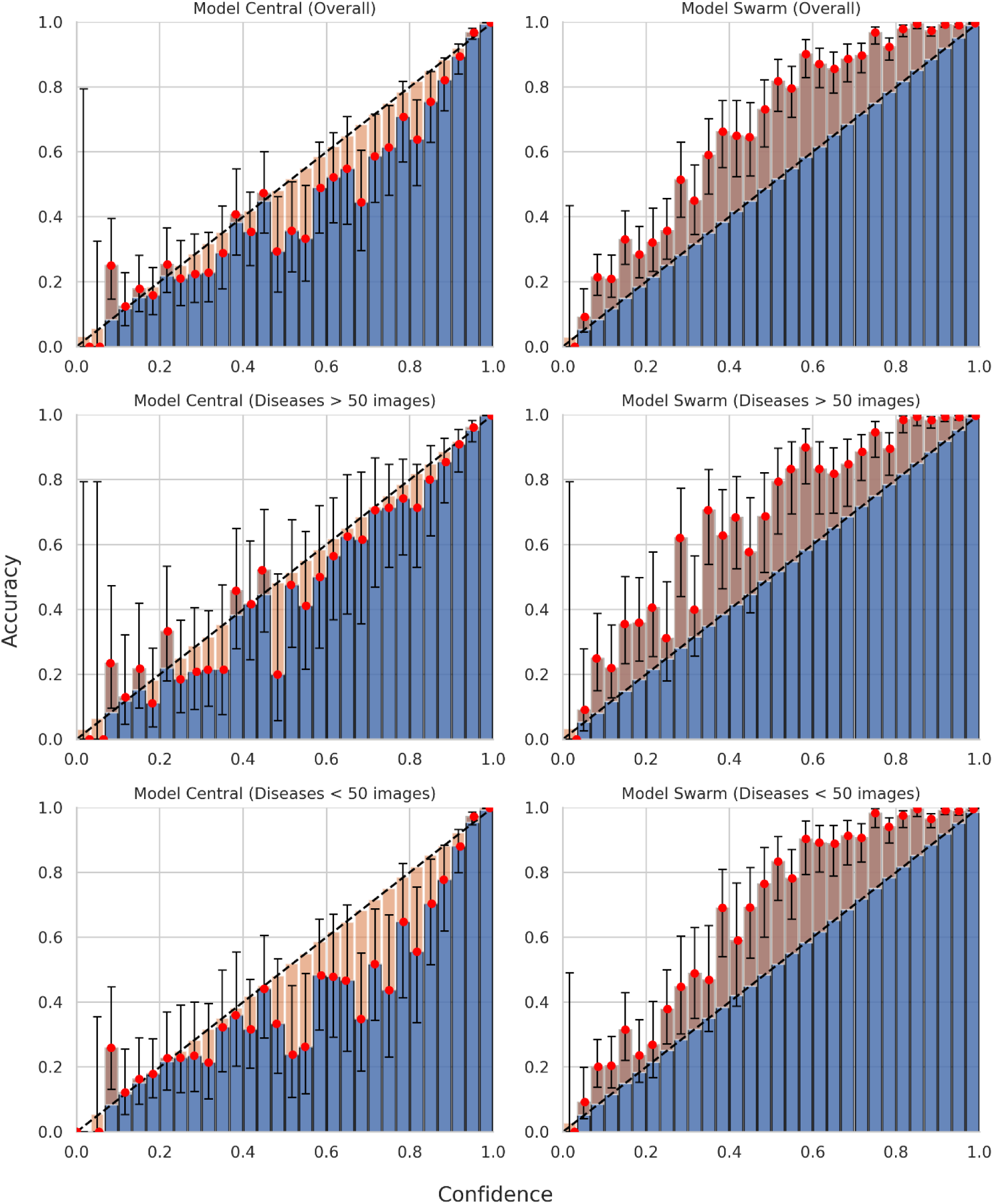
Expected Calibration Error (ECE) plots for the Central and Swarm models. The x-axis represents confidence scores divided into 30 bins, while the y-axis shows the corresponding accuracy. The dotted black line indicates the ideal calibration line. The top panel presents overall performance of the Central and Swarm models. Diseases are further categorized into high-representation (>50 images) and low-representation (<50 images) groups, with the corresponding performances of the Central and Swarm models shown in the middle and bottom panels, respectively.

We also quantified epistemic uncertainty that arises from limited knowledge or insufficient training data, using the BALD (Bayesian Active Learning by Disagreement) metric, computed on a per-class basis (Figure S5). BALD captures the degree to which model predictions vary under different plausible model parameter configurations and therefore serves as an indicator of model robustness to parameter-level uncertainty.

The BALD metric was computed using multiple stochastic forward passes through the Centrally and Swarm trained ResNet-100 model, implemented via Monte Carlo Dropout at inference time. For each input image, predictive probabilities were sampled repeatedly to estimate the distribution of outputs under different model parameter realizations. BALD was then calculated as the difference between the total predictive entropy and the expected entropy across stochastic passes, thereby isolating epistemic uncertainty.

The Centralized and Swarm models exhibit broadly similar BALD distributions; however, the Swarm model displays lower BALD values for several underrepresented classes, most notably for the classes with the smallest sample sizes (class indices >175). This reduction in epistemic uncertainty indicates that swarm learning fosters more stable and generalizable feature representations, particularly for low-prevalence disorders, likely due to the decentralized aggregation of heterogeneous data sources.

### Advancing next generation phenotyping through Swarm learning

We next investigated the performance gains achieved through Swarm training relative to the current state-of-the-art GestaltMatcher algorithm. The existing GestaltMatcher framework supports 275 disorders, whereas the proposed swarm model in this study, jointly trained on the GMDB and AIDY datasets, supports 338 disorders. A complete list of supported disorders is provided in Table S6.

For this comparison, we evaluated the ResNet-100–based GMDB model against the swarm learning model, as the ResNet-100 architecture demonstrated superior performance compared to the ResNet-50 model. Model performance was assessed across demographic subgroups, including age (baby, child, and adult), sex (male and female), and ethnicity (Asian, African, and Caucasian). The results are presented in Figure 4A.

**Figure 4.**
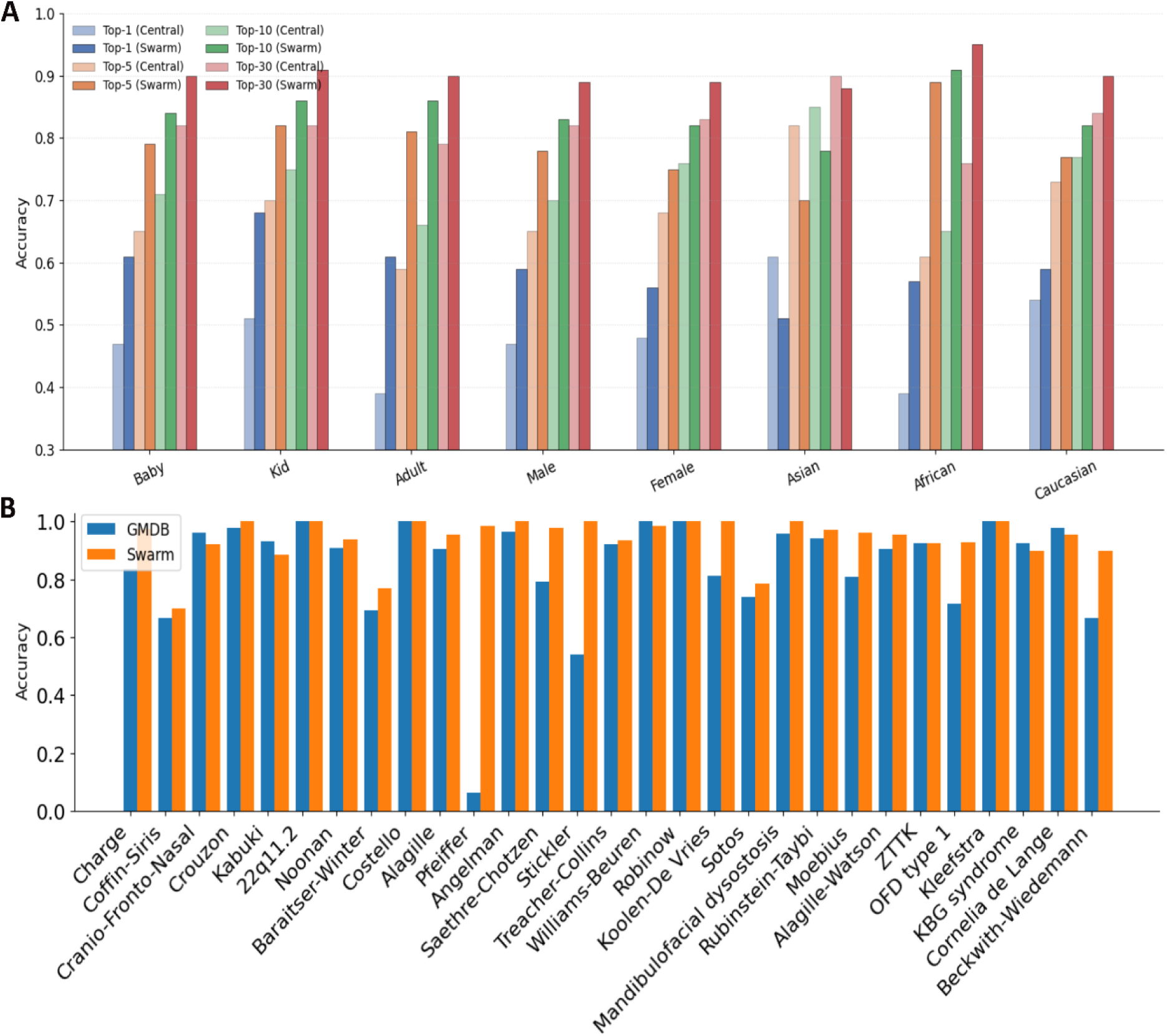
A) Performance comparison between GestaltMatcher model and current advance Swarm model across age (baby, child, and adult), gender (male and female), and ethnicity (Asian, African, and Caucasian) for all Top-K. B) Comparison of GestaltMatcher model and Swarm model performances for some selected clinically challenging disorders is shown for Top 10 evaluation.

Across all age groups, the swarm model demonstrated substantial performance improvements over the GMDB model across all Top-K metrics, with gains ranging from 7–25%. The improvement was particularly pronounced in the adult cohort. This observation is likely attributable to the fact that the GMDB dataset and the original GestaltMatcher framework were predominantly trained on infant and pediatric facial images, resulting in limited representation of adult phenotypes. In contrast, the AIDY dataset contributes a larger proportion of mature facial images, enabling the swarm model to benefit from both developmental facial representations from GMDB and mature facial representations from AIDY. Consequently, the swarm model exhibited consistently superior performance across all age groups.

Similarly, performance improvements were observed across sex-based subgroups. Although male and female samples were approximately balanced in both datasets, the swarm model consistently outperformed the original GestaltMatcher model across all evaluated Top-K metrics. The improved performance of the swarm model is likely attributable to the increased phenotypic diversity and cross-institutional heterogeneity introduced through decentralized Swarm training. Although the datasets were approximately balanced by sex, swarm learning enabled exposure to a broader distribution of facial phenotypes, imaging conditions, and syndrome-specific variations. This results in a more robust and generalized feature representations. This enhanced representation learning likely contributed to improved discrimination across both sex-based subgroups. Similarly, with the exception of the Asian subgroup, the Swarm model demonstrated superior performance in both the African and Caucasian subgroups.

In Figure 4B, we compare GMDB model with Swarm model across several clinically challenging disorders that exhibit phenotypic overlap, and are therefore difficult to diagnose, even for experienced clinicians. Several disorders in this cohort belong to related molecular or phenotypic groups. For example, Noonan syndrome and Costello syndrome are part of the RASopathy spectrum, while Pfeiffer syndrome, Saethre-Chotzen syndrome, and Crouzon syndrome are syndromic craniosynostosis disorders with overlapping craniofacial morphology.

Other syndromes, such as 22q11.2 deletion syndrome, Williams-Beuren syndrome, and Angelman syndrome, demonstrate marked phenotypic variability across affected individuals. Many of these diseases also show variable expressivity, age-dependent facial changes, and limited case representation in clinical datasets. We found that for most of these disorders, the Swarm model outperformed the current GestaltMatcher algorithm. This suggests an enhanced ability of the Swarm model to generalize subtle phenotypic patterns across distributed datasets while preserving disease-specific facial characteristics relevant for clinical recognition. Such improvements may be particularly important for underrepresented rare diseases, where data heterogeneity and small cohort sizes can negatively affect centralized model training. A complete list of disorders currently supported by the Swarm model is provided in Table S6.

## Conclusions

Swarm learning possesses transformative potential in distributed machine learning that facilitate edge computing, blockchain-based peer-to-peer networking, and decentralized coordination to enable collaborative model training without centralizing sensitive data (23,24,28,29). It allows administratively independent nodes to train models locally on their proprietary data while only share model parameters. This mechanism achieved superior performance compared to locally trained models. Initially, with COVID-19 in transcriptomic data, it has been demonstrated that the swarm model performs better than or similar performance to individual nodes. These results have been demonstrated on swarms with up to six nodes with different prevalences, even different techniques for data acquisition at the nodes. Further, using a popular, openly available on Kaggle, X-ray chest image dataset, Swarm learning was used to successfully classify different lung diseases (30). Swarm learning has also been successfully applied in other various medical contexts (25–27,31–35).

While swarm learning has demonstrated technical feasibility and superiority over local training in several medical domains, its application to rare disease diagnosis remains limited. The paradox is significant as swarm learning explicitly promises to overcome data scarcity challenges through distributed training, yet real-world deployments have remained sparse. This gap reflects a constellation of unresolved challenges between conceptual potential and practical implementation. Furthermore, in previous Swarm trained models were only limited to a few very well-known and well-categorized diseases. On that capacity, craniofacial disorders represent a unique and compelling context where swarm learning could theoretically deliver transformative clinical value. A primary barrier to rare disease deployment in swarm learning is the compounded class imbalance problem that emerges at multiple levels. Each participating institution with low disease prevalence may have zero, one, or a handful of cases of a rare disease. This creates sparse signal at the node level, making local model training noisy and unreliable. When each node trains separately on severely imbalanced data, then aggregates parameters through blockchain-based consensus mechanisms, the learned representations may not adequately capture the rare disorder phenotype. A node with zero disease cases learns only control representations; its aggregated parameters dilute disease-specific features learned by other nodes. Amid this, we would like to investigate how well swarm model generalizes to rare diseases.

We considered two complementary datasets, GMDB, characterized by broad syndromic and patient diversity, and AIDY, containing a larger number of images per patient and more mature facial phenotypes. Considering these datasets, we showed that distributed model training can preserve diagnostic performance while maintaining strict data locality and institutional autonomy. Across a comprehensive evaluation framework that included in-distribution, cross-institutional, and rare-disorder assessments, swarm-trained models consistently matched the accuracy of centrally trained models and outperformed institution-specific local models, particularly in scenarios with substantial domain shift. Importantly, our analyses reveal that decentralized training does not erode sensitivity to low-prevalence, ultra-rare syndromes despite the inherent data fragmentation across sites. Performance for rare disorders remained comparable between local, centralized, and swarm models, indicating that the parameter-aggregation mechanisms underlying swarm learning are sufficiently robust to retain salient features associated with underrepresented phenotypes. This is further supported by our uncertainty quantification results. The swarm model exhibited a more conservative calibration profile, with empirical accuracies exceeding predicted confidences across confidence bins. This represents reduced risk of high-confidence misclassification, a critical requirement for clinical deployment. Furthermore, swarm learning yielded lower epistemic uncertainty for several of the smallest classes, suggesting improved feature stability and generalizability for rare disorders derived from the integration of heterogeneous, non-pooled training distributions.

Beyond technical performance, our work addresses key structural challenges that currently limit the scalability and inclusivity of NGP. Centralized training pipelines require aggregation of sensitive patient images to a single repository. This is an expensive approach and incompatible with modern privacy regulations, cross-border data governance, and institutional data sovereignty. Swarm learning circumvents these constraints by enabling collaborative model training without sharing raw data, using blockchain-mediated coordination to ensure secure, transparent, and fault-tolerant parameter synchronization. This paradigm not only preserves patient privacy but also democratizes model development by enabling participation from institutions with diverse populations, imaging protocols, and disease spectra. These factors are essential for improving performance in global clinical settings.

Our results demonstrate that decentralized swarm learning is a powerful and scalable framework for training deep facial phenotyping models in rare disease diagnostics. It facilitates the integration of geographically and demographically diverse datasets without compromising privacy. It maintains diagnostic accuracy comparable to centralized training with improved calibration and uncertainty characteristics. Looking ahead, this strengths position swarm learning not merely as a privacy-preserving alternative to centralized training, but as a foundational infrastructure for the future rare disease diagnostics. Rather than diluting disease-specific signals, swarm models retain sensitivity despite severe data scarcity across institutions. This directly addresses the most persistent challenges in rare disease diagnosis. This ability together with robust uncertainty calibration make swarm-model safer choice for clinicians that reduces overconfident misclassifications and provides clinicians more trustworthy confidence estimates, a critical requirement for decision support in genetic diagnosis. In the longer term, integrating swarm learning with multimodal signals, such as genomic variants, longitudinal clinical records, and other imaging modalities etc., could enable richer phenotype–genotype discovery while minimizing data exposure risks. As such models are integrated into clinical workflows, they may facilitate earlier recognition of rare genetic disorders. This will shorten ‘diagnostic-odyssey’ and improve referral prioritization.

## Materials and Methods

### GMDB data

GestaltMatcher Database (GMDB, https://db.gestaltmatcher.org/photos?blank=true) collects medical images of individuals with clinically or molecularly confirmed diagnoses from publications and individuals that gave appropriate consent for the research. It is open to clinicians and researchers working in medical research fields. It’s a service operated by the <u>Association for Genome Diagnostics (AGD)</u>, which is a registered non-for-profit organization in Germany. Access of the data is controlled by a reviewing committee to avoid data abuse. GMDB (v1.1.0) contains 10,980 frontal images of 8,346 patients with 576 different disorders. All the disorders have at least two patients. The dataset was further divided into two sets, a “frequent” (GMDB-Frequent) and a “rare” (GMDB-Rare) set. The disorders with more than six patients were assigned to GMDB-Frequent, while the disorders with six or fewer patients were assigned to GMDB-Rare. There are 9,693 images of 7,314 patients with 277 disorders in GMDB-Frequent and 1,287 images of 1,032 patients with 299 disorders in GMDB-Rare.

### AIDY data

We are starting with a dataset of 30,000 patients and over 1 000 000 images from photographic archives of the Maxillofacial Surgery, Plastic Surgery, and Medical Genetics departments at Necker–Enfants Malades Hospital (AP-HP). This database, created by the Departments of Craniomaxillofacial Surgery and Medical Genetics, was made possible by the efforts of a dedicated hospital photographer. Using a professional studio and adhering to strict consent protocols, the database was developed with a focus on ethical standards and privacy. Clinical Research Associates (CRAs), along with ergonomic tools, were used to confirm genetic diagnoses for each patient and selecting qualified images from the database for high-quality data curation. Codoc (Dr. Warehouse) will be used for local patient selection, and once patients are identified, their associated images will be automatically retrieved. To expedite the curation process, we have identified RetinaFace, an algorithm for detecting facial landmarks (eyes, mouth, nose), and 6DRepNet360, which analyzes facial orientation in photographs. These algorithms will assist in pre-annotating images (e.g., detecting if photos contain 0, 1, or multiple faces, and identifying frontal or profile views). CRAs will then correct the pre-annotations and further assess image quality.

The patient inclusion criteria are:

- Patients followed in medical genetics,
- Patients undergoing maxillofacial surgery, or craniofacial surgery as part of the management of a pathology, of genetic origin or not, associated with dysmorphism of the head and neck,
- Patients for whom frontal and profile facial photographs are taken as part of their treatment.

The inclusion criteria for control subjects are:

- Patients followed in maxillofacial surgery, for a disease other than a rare disease associated with dysmorphia in the head or neck: acute pathology (wound) or chronic (gynecomastia).
- Patients for whom frontal and profile facial photographs are taken as part of their treatment.

The criteria for non-inclusion of patients are:

- Patients who have undergone facial or skull surgery before the first photo was taken.
- Person subject to a judicial safeguard measure.
- People objecting to the reuse of their health data. The criteria for non-inclusion of control subjects are:
- Pathologies affecting facial symmetry (dental cellulitis, displaced fractures).
- Patient followed for dysmorphic syndrome or in whom dysmorphic syndrome has been suspected.
- Person subject to a judicial safeguard measure.
- People objecting to the reuse of their health data.

For this work total 30,569 images were considered from 3,540 patients with 351 disorders. Following definition of GMDB Frequent and Rare, this comprises of 26,237 and 4,332 images from 2,967, 589 patients for Frequent and Rare disorders.

To ensure consistency across datasets and facilitate reliable mapping and integration, syndromes have been named following a standardized hierarchy. Whenever possible, the Orphanet (ORPHA) code was used as the primary reference due to its specificity and recognition in rare disease classification. When no ORPHA code was available, the OMIM (Online Mendelian Inheritance in Man) identifier was used as an alternative. In some cases, where no clear syndrome designation could be found in either ORPHA or OMIM, the syndrome was labeled using the associated gene name (e.g., “CDH8-related disorder”). This approach ensures that each entry remains identifiable and traceable, even in the absence of an established clinical syndrome name.

### Deep Learning Model

A widely used variant of the ResNet architecture, known as iResNet, with varying network depths, was employed (23). Pre-trained models supplied by insightface (https://github.com/deepinsight/insightface) that have been trained on Glint360k face recognition datasets (39) using Additive Angular Margin Loss (Arc-Face) (40) with iResNet architecture (41) of depth 50 and 100 were used. The loss is defined as:

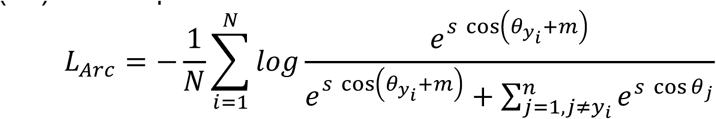

where 𝜃_𝑗_ is the angle between weights 𝑊_𝑗_ and feature 𝑥_𝑖_. Angular margin 𝑚 was inserted, such that the angle between the true logit, 𝑦_𝑖_, and the representation vector becomes 𝜃_𝑦𝑖_ +𝑚. 𝑠 is the scale of 𝐿_2_ normalized representation vectors. The pre-trained models used 𝑚 and 𝑠 set to 0.5 and 64. Representations learned using this loss exhibited stronger separation between different classes and greater similarity within the same class compared to most other metric learning losses (23). These models can be downloaded from: https://github.com/deepinsight/insightface/tree/master/model_zoo. An important preprocessing step of insightface’s training procedure is to align faces based on five landmarks: left and right eye, nose, left and right mouth corner. For our implementation, we used RetinaFace (42) to obtain these land-marks and the alignment code they supplied, which uses an affine transformation based on matching landmark locations. Additionally, minor adjustments to the model architecture were added. We removed the batch normalization of the computed features. This normalization was necessary for ArcFace, which we did not use during fine-tuning. Instead, due to the small dataset size, significant class imbalance, and long tail distributions, we decided only to optimize Weighted Cross Entropy Softmax Loss (WCE). Model was fine-tuned on GMDB data using aligned faces of size 112×112, randomly flipping horizontally, randomly converting color images to gray, color jittering, and randomly adding zooming/cropping artifacts. Adam optimizer with a base learning rate of 1e-3 was used with reduction by a factor of 2 when the top-5 mean accuracy on the validation set plateaus until convergence.

For inference, model ensemble and test time augmentation was used to improve the performance on both seen and unseen disorders by computing multiple representation vectors per image, aiming to end up with a better overall ranking than each separate representation vector. One face verification model, one deeper disorder model (using iResNet-100), and one disorder model (using iResNet-50) were used. During Test time augmentation (TTA), an image and augmented versions of that image (e.g., horizontally flipped, converted from color to gray, rotation, and translation) were used. Finally, the cosine distance of all models in the ensemble and TTAs per model (i.e., three models and two TTA each, 3×2×2=12 cosine distances) were averaged. The order of the disorders ranked for verification was determined by the k-nearest neighbours of the average cosine distance between the gallery images to the test image.

### Swarm setup

Swarm Learning is a decentralized learning designed for privacy-preserving collaborative machine learning. It is developed by Hewlett Packard Enterprise (HPE) https://github.com/HewlettPackard/swarm-learning. It eliminates the need for a central leader by leveraging blockchain, edge computing, and distributed AI principles. The architecture includes the following core components:

#### Swarm Learning (SL) Nodes

Each SL node operates in a modular container and interfaces with the ML model (hosted by an ML node). The SL node manages model parameter synchronization, securely sharing and aggregating learnings with other SL nodes in the network. SL nodes synchronize periodically, governed by a sync frequency, which specifies the number of local training batches completed before communication. <u>Swarm Network (SN)</u> <u>Nodes:</u> SN nodes form a blockchain network to manage state and coordinate activities across SL nodes. Uses an open-source Ethereum blockchain to store metadata, such as synchronization history, while ensuring immutability and security. One SN node is designated as the Sentinel Node, responsible for initializing the blockchain. <u>ML Nodes:</u> ML nodes train machine learning models using local, private data. Each ML node pairs with an SL node to integrate the locally trained model into the decentralized framework. <u>Swarm Operator (SWOP)</u> <u>Nodes:</u> SWOP nodes handle task orchestration, such as initiating training runs, container management, and model sharing. They interact with Taskrunner Frameworks for decentralized task management, using YAML profiles to configure resources, SL nodes, and policies. <u>Swarm Command Interface (SWCI) Nodes:</u> Provides a command-line interface for managing and monitoring the Swarm Learning framework. SWCI nodes can issue commands like creating contexts, managing tasks, and retrieving node statuses. <u>License Server:</u> Ensures that the necessary licenses are available for operating the Swarm Learning framework, managed via an AutoPass License Server (APLS).

All communications between nodes are secured using X.509 certificates. Blockchain ensures that no raw data is shared. Only aggregated model updates are exchanged. All nodes, including SL, SN, ML, and SWOP, are deployed as Docker containers for modularity and scalability. Each container operates independently, ensuring secure and isolated functionality.

Swarm Learning nodes interact through a coordinated workflow to securely train and synchronize machine learning models. SL nodes communicate with their paired ML nodes via named pipes (FIFO) for exchanging model parameters and training updates. Use a file server (default port: 30305) to share model parameters securely with other SL nodes. Regularly register their state with SN nodes through the SN API port (default: 30304). SN nodes use a peer-to-peer (P2P) blockchain port (default: 30303) to exchange blockchain state information with other SN nodes. Metadata regarding synchronization events and node states is logged in the blockchain. SWOP nodes execute tasks such as starting and stopping Swarm runs or managing containers. SWCI nodes interact with SN nodes using APIs to provide real-time monitoring and management of the Swarm Learning framework. Sync Frequency governs how often nodes share updates. A large sync frequency reduces communication overhead, while a small sync frequency ensures frequent updates but slows down training. Adaptive sync frequency adjusts the interval dynamically based on model performance metrics, such as the mean loss.

If a node (e.g., SL or SN) fails, the network can dynamically adjust by reassigning leadership or redistributing tasks. For example: Leader Failure Detection and Recovery (LFDR) allows SL nodes to continue synchronization even if the designated leader node fails.

The Swarm Learning framework employs a distributed model parameter merging process to aggregate updates from all SL nodes securely and effectively. After completing a sync frequency, all SL nodes transmit their local model parameters to a designated leader node. The leader aggregates the parameters using one of three user-defined methods:

Mean (Default), Coordmedian (Computes the coordinate-wise weighted median of parameters, which is robust to outliers), Geomedian (Uses Weiszfeld’s algorithm to calculate the geometric median, iteratively refining the parameter set to minimize deviation).

Parameter Redistribution: The aggregated model parameters are redistributed to all SL nodes for the next round of training.

Node Weightage: Users can assign relative importance to individual nodes (e.g., based on data quality or size) by configuring a weightage between 1 and 100.

Minimum Peers: The number of SL nodes required to complete synchronization is configurable, ensuring robust collaboration.

Adaptive Synchronization: The framework dynamically adjusts the sync frequency by monitoring the model’s mean loss:If training progress is steady, the sync interval is increased to improve efficiency. If progress stalls, synchronization occurs more frequently to refine updates.

All swarm nodes were initialized with an identical base model architecture, optimizer configuration, and training protocol to ensure consistency across sites. A private, permissioned blockchain network was established to coordinate decentralized learning, model parameter exchange, and node authentication. Each center was assigned a unique cryptographic identity for secure participation in the swarm, and access controls were enforced through smart contracts governing node registration and training rounds. Local training was performed independently at each center using its site-specific dataset. After a predefined number of local training iterations, model weight updates were encrypted and shared with the swarm network. A decentralized aggregation protocol was then executed to compute a global model update without exposing individual center contributions. The aggregated global model was redistributed to all participating nodes, where it served as the initialization for the subsequent training round. This iterative process of local training, secure parameter sharing, and decentralized aggregation continued until desired criteria reached.

Network communication was synchronized using secure message passing with fault-tolerant mechanisms to allow continued training in the presence of transient node delays. Comprehensive logging was implemented at each node to track training progress, synchronization status, and aggregation events for reproducibility and auditability.

## Ethics

This research was approved by the Institutional Review Board of GMDB and AIDY.

## Data Availability

The data used in this study are not available for public access due to hospital confidentiality agreements, in compliance with data protection regulations.

## Supplementary information

**Table S1.**
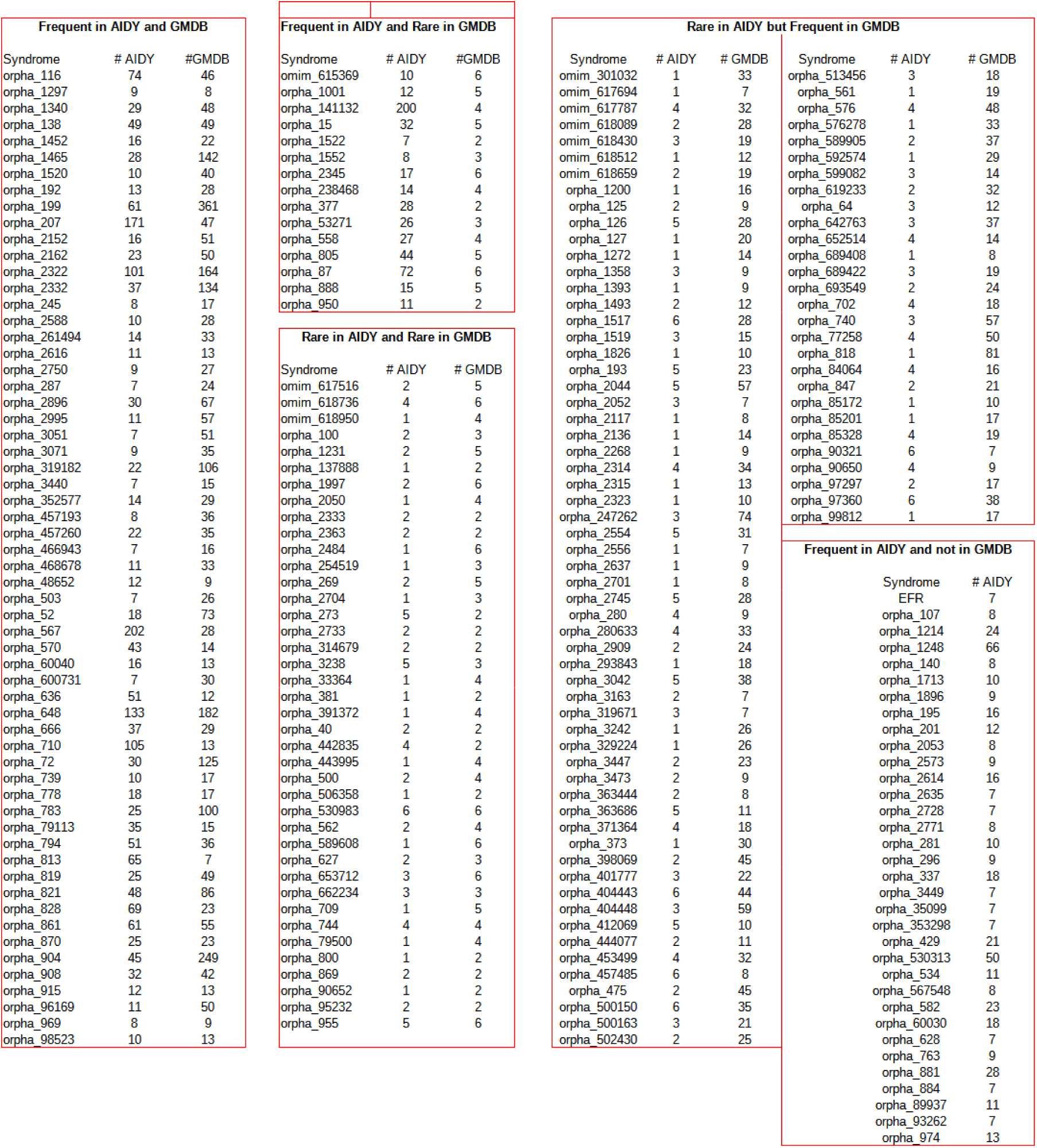
Complementarity of the datasets.

**Figure S1.**
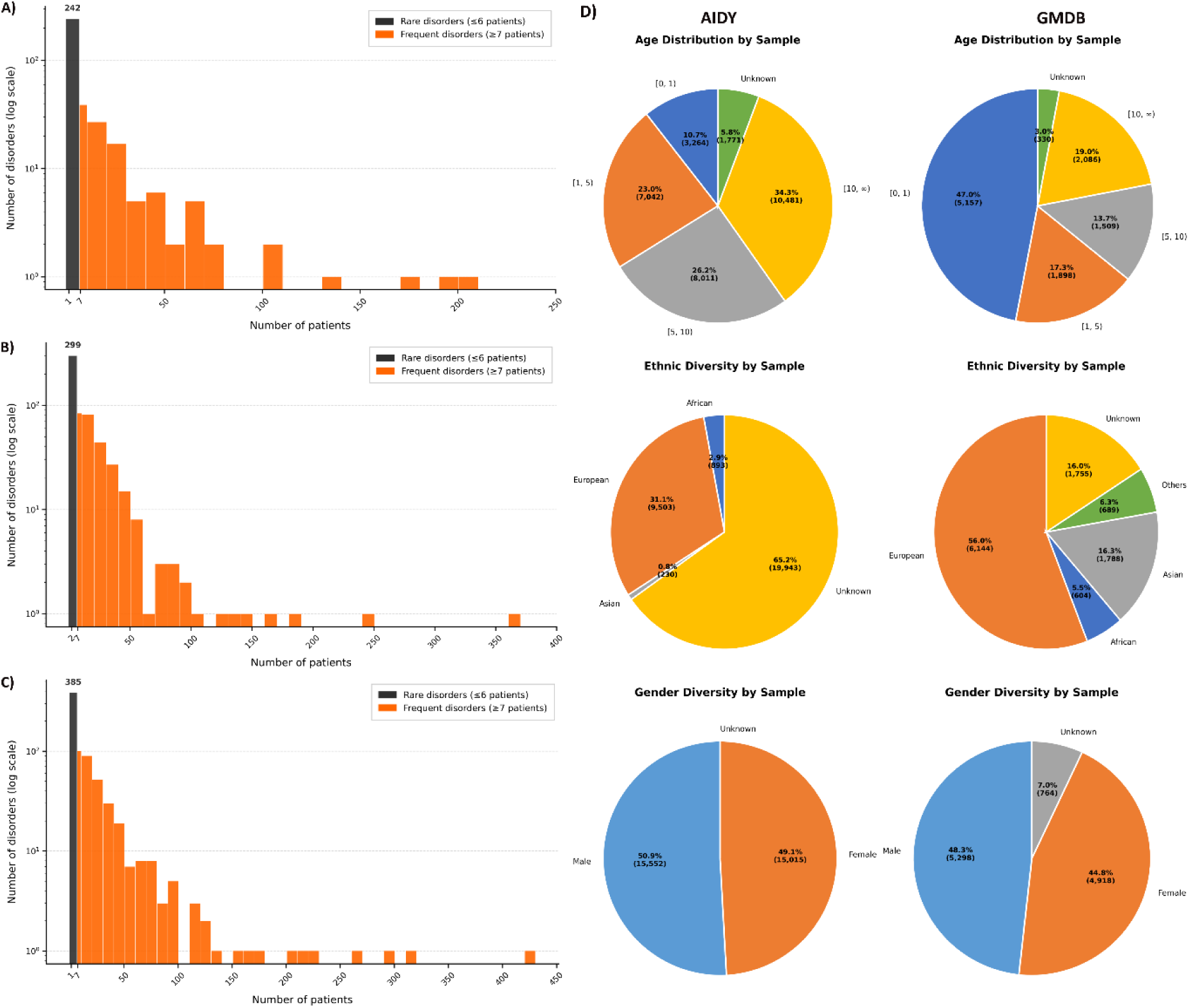
Disorder distribution in AIDY (A), GMDB (B) and together (C) datasets. Black bar represents number of rare disorders with < 7 patients and orange bar represents frequent disorders with > 6 patients. Demographic diversity of GMDB and AIDY datasets is shown in D. Age, Ethnicity, and Sex are compared.

**Figure S2.**
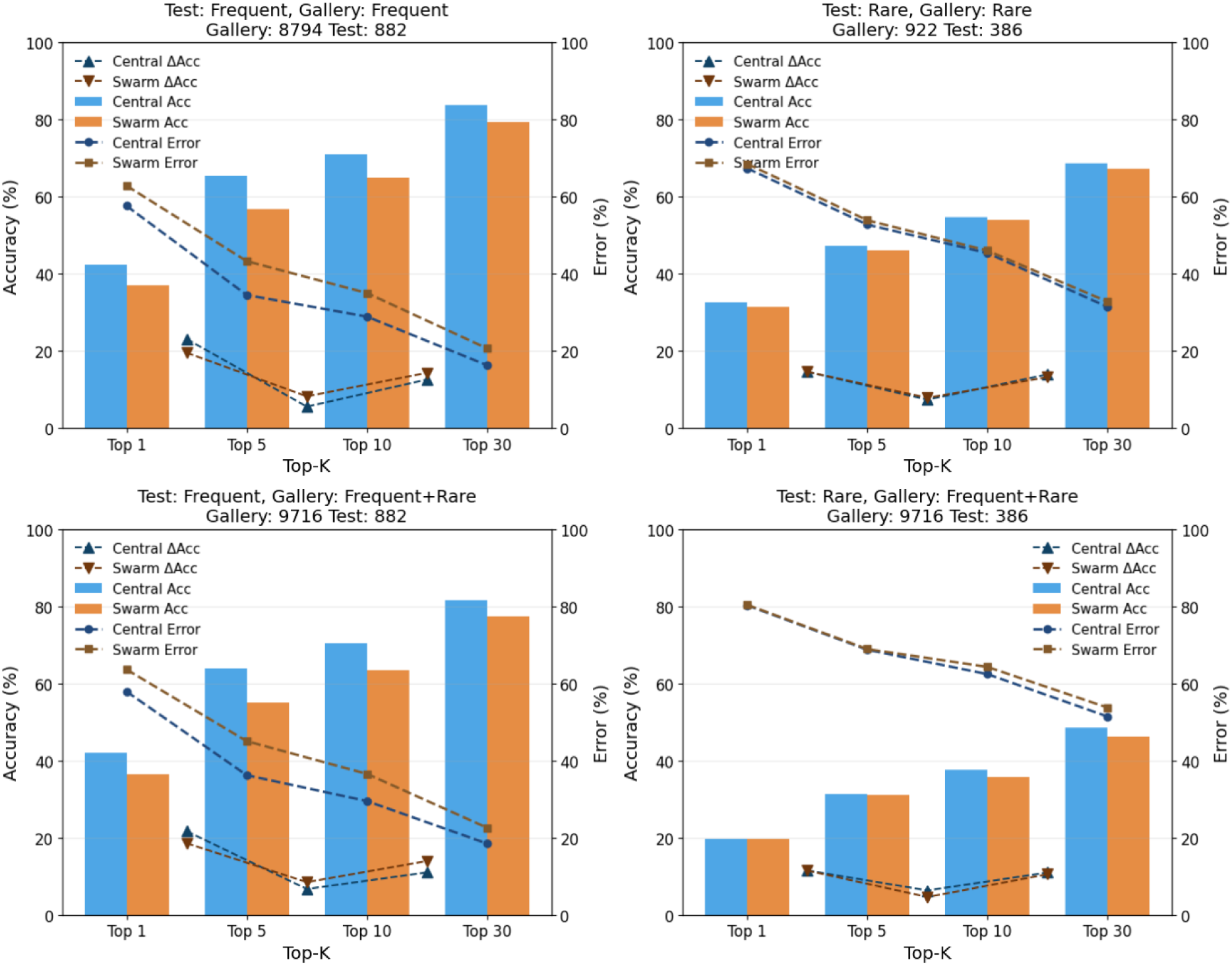
Comparison of accuracy of Swarm (2 nodes) and Central model trained on GMDB data. Model ensemble and TTA method were used for model evaluation. Top 1, Top 5, Top 10 and Top 30 matrices were shown for various data split, mentioned at top. Dotted square and circle represents Error in %. Dotted triangle represents gain in accuracy between Top-K.

**Figure S3.**
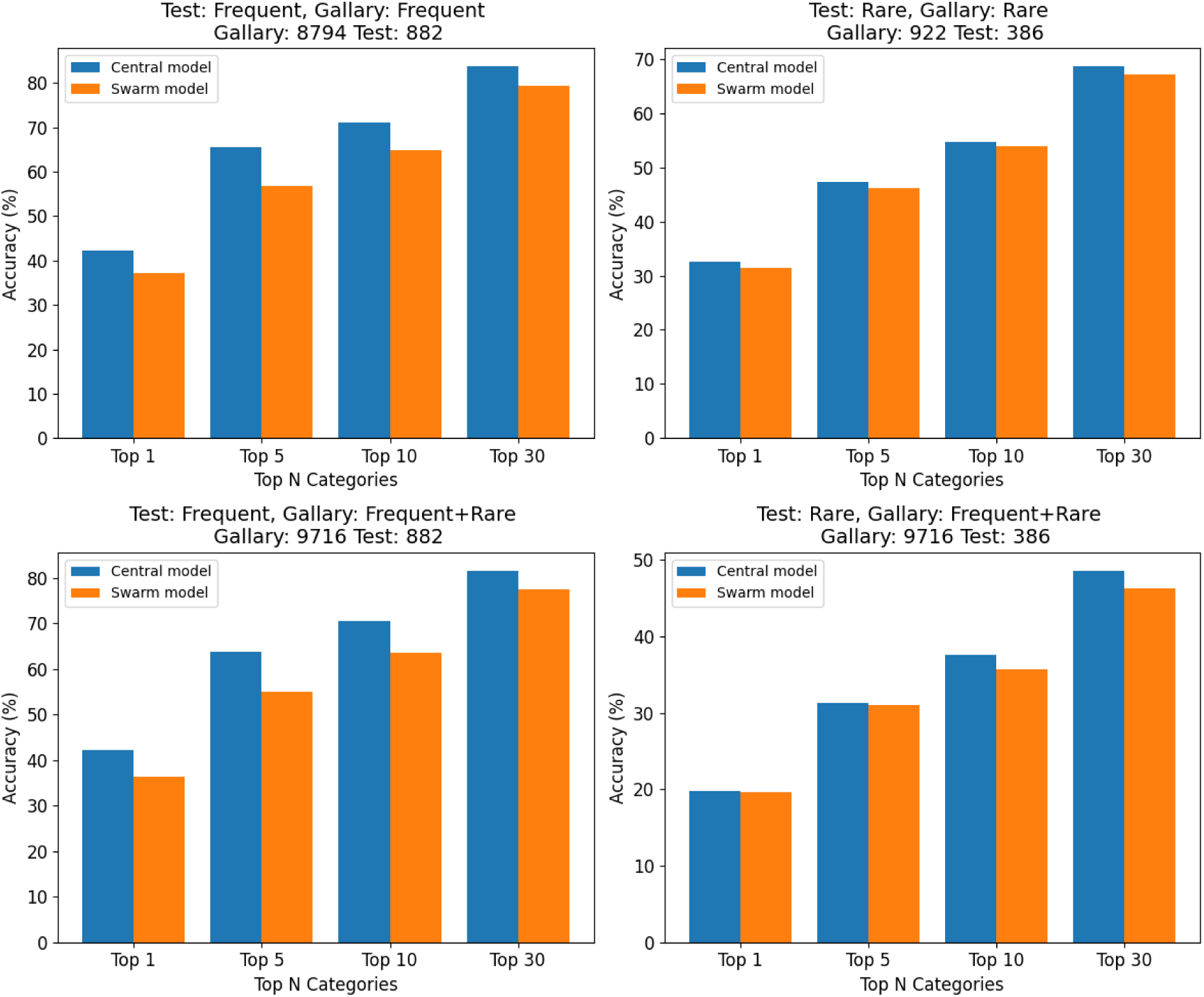
Performance comparison of swarm and central models under a sex-based non-stratified split into two nodes.

**Figure S4.**
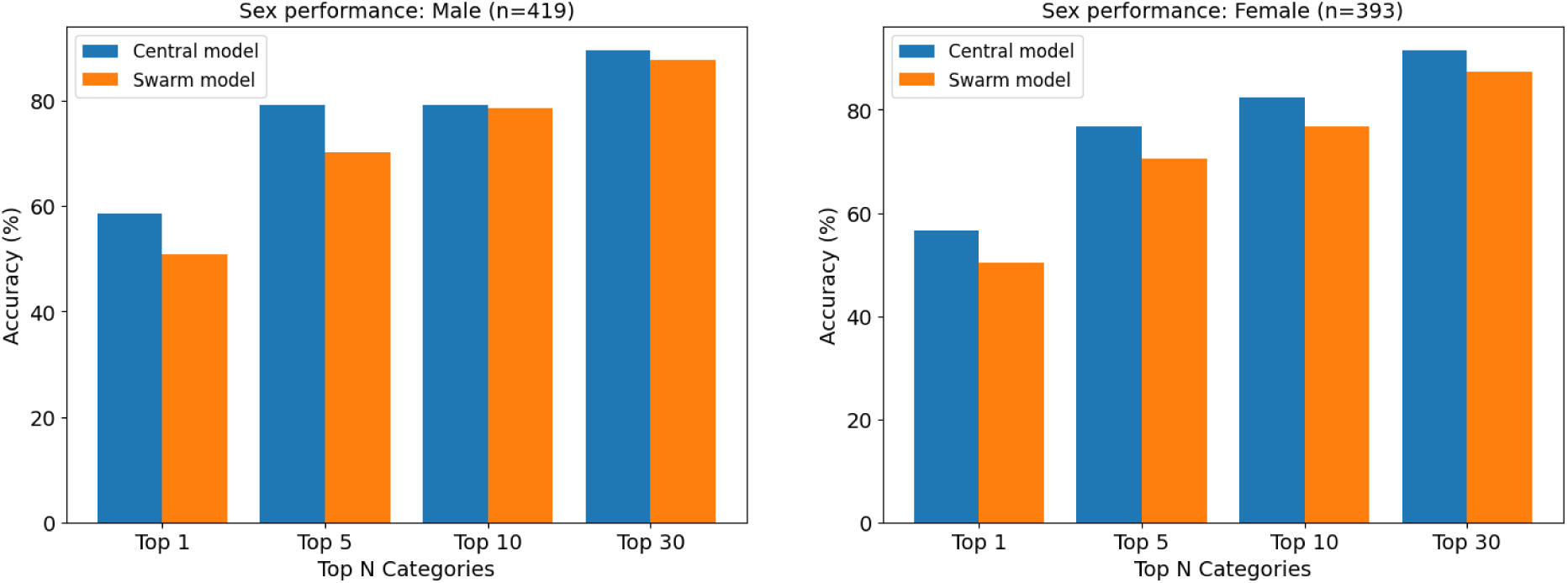
Swarm model performance on male and female test data under a sex-based non-stratified split into two nodes.

**Figure S5.**
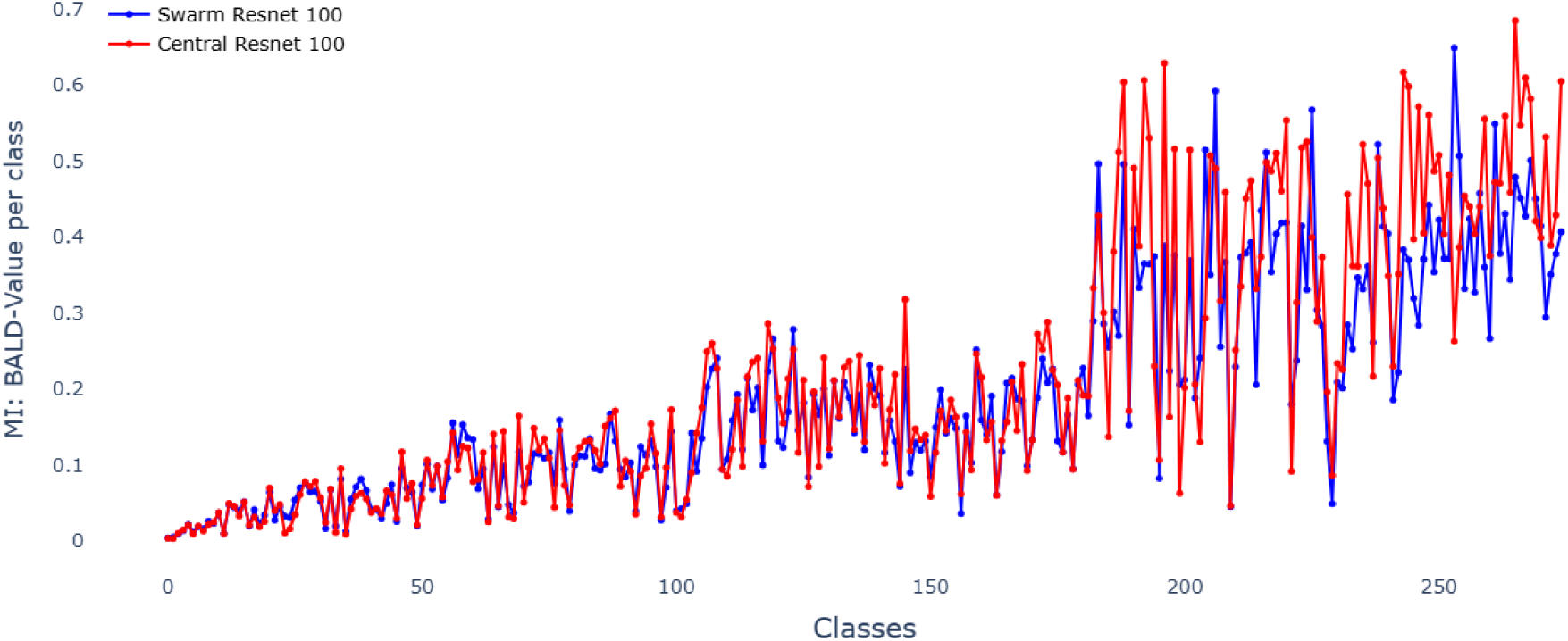
Bald epistemic uncertainty for the ResNet-100 model across individual disease classes, comparing Centrally trained (red) and Swarm-trained (blue) models.

**Table S2.** Performance of GMDB model.

| On GMDB test data (with GMDB gallery) |  |  |  |  |
| --- | --- | --- | --- | --- |
| Category | Top 1 | Top 5 | Top 10 | Top 30 |
| Per Syndrome Mean: Frequent/Frequent | 41.23 | 64.43 | 70.46 | 83.66 |
| Per Syndrome Mean: Rare/Rare | 32.5 | 47.42 | 54.79 | 69.54 |
| Per Syndrome Mean: Frequent/Overall | 40.97 | 63.17 | 68.59 | 80.58 |
| Per Syndrome Mean: Rare/Overall | 19.71 | 31.74 | 37.71 | 47.6 |
| On AIDY test data with GMDB gallery |  |  |  |  |
| Category | Top 1 | Top 5 | Top 10 | Top 30 |
| Per Syndrome Mean: Frequent/Frequent | 33.22 | 54.52 | 61.76 | 80.43 |
| Per Syndrome Mean: Rare/Rare | 15.81 | 30.67 | 41.28 | 55.25 |
| Per Syndrome Mean: Frequent/Overall | 30.61 | 53.58 | 59.31 | 75.05 |
| Per Syndrome Mean: Rare/Overall | 6.97 | 18.3 | 20.74 | 36.54 |

**Table S3.**
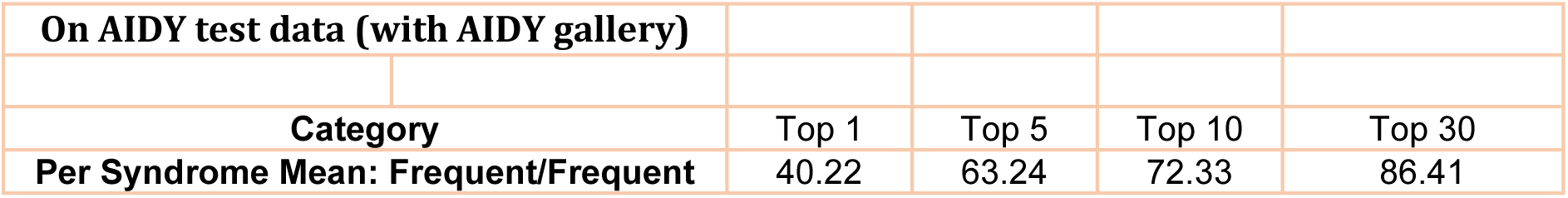
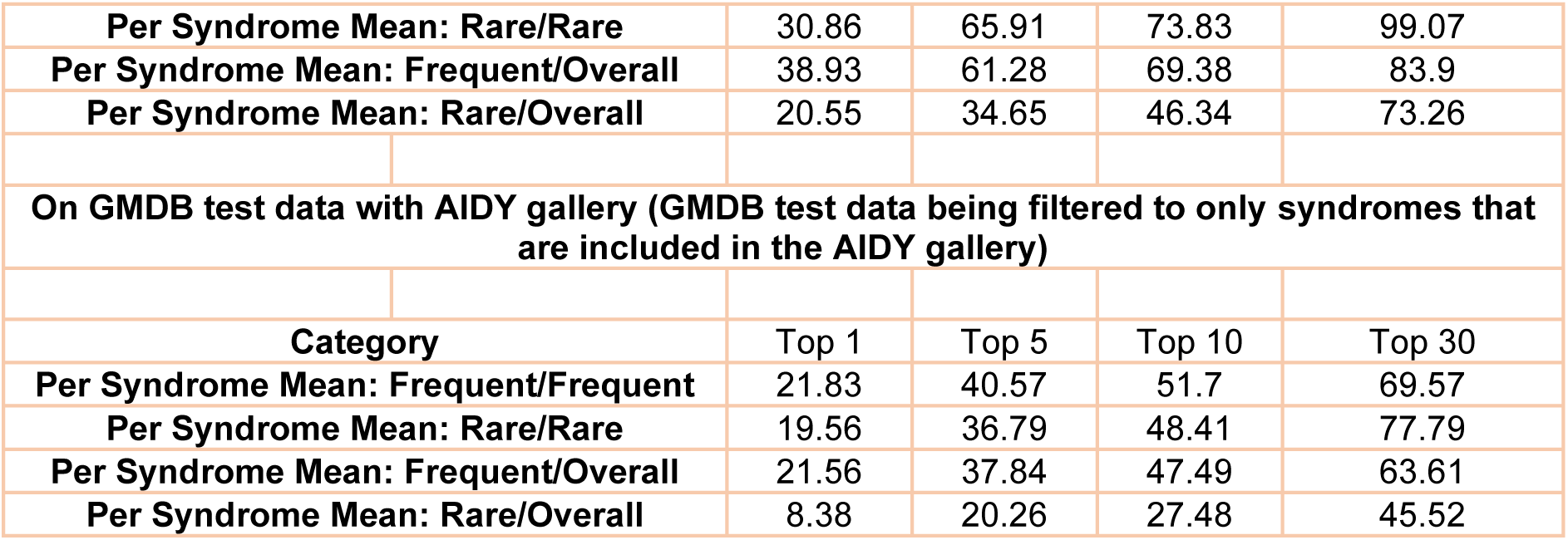
Performance of AIDY model.

| On AIDY test data (with AIDY gallery) |  |  |  |  |
| --- | --- | --- | --- | --- |
| Category | Top 1 | Top 5 | Top 10 | Top 30 |
| Per Syndrome Mean: Frequent/Frequent | 40.22 | 63.24 | 72.33 | 86.41 |
| Per Syndrome Mean: Rare/Rare | 30.86 | 65.91 | 73.83 | 99.07 |
| Per Syndrome Mean: Frequent/Overall | 38.93 | 61.28 | 69.38 | 83.9 |
| Per Syndrome Mean: Rare/Overall | 20.55 | 34.65 | 46.34 | 73.26 |
| On GMDB test data with AIDY gallery (GMDB test data being filtered to only syndromes that are included in the AIDY gallery) |  |  |  |  |
| Category | Top 1 | Top 5 | Top 10 | Top 30 |
| Per Syndrome Mean: Frequent/Frequent | 21.83 | 40.57 | 51.7 | 69.57 |
| Per Syndrome Mean: Rare/Rare | 19.56 | 36.79 | 48.41 | 77.79 |
| Per Syndrome Mean: Frequent/Overall | 21.56 | 37.84 | 47.49 | 63.61 |
| Per Syndrome Mean: Rare/Overall | 8.38 | 20.26 | 27.48 | 45.52 |

**Table S4.** Performance of Centralized model.

|  |  |  |  |  |
| --- | --- | --- | --- | --- |
| On GMDB test data (with GMDB gallery) |  |  |  |  |
| Category | Top 1 | Top 5 | Top 10 | Top 30 |
| Per Syndrome Mean: Frequent/Frequent | 43.04 | 62.77 | 70.02 | 81 |
| Per Syndrome Mean: Rare/Rare | 31.3 | 48.65 | 55.94 | 69.43 |
| Per Syndrome Mean: Frequent/Overall | 42.05 | 62.2 | 69.45 | 79.11 |
| Per Syndrome Mean: Rare/Overall | 19.13 | 31.23 | 36.92 | 47.74 |
| On GMDB test data with both GMDB and AIDY gallery |  |  |  |  |
| Category | Top 1 | Top 5 | Top 10 | Top 30 |
| Per Syndrome Mean: Frequent/Frequent | 41.17 | 61.51 | 70.45 | 81.36 |
| Per Syndrome Mean: Rare/Rare | 29.49 | 46.02 | 54.57 | 68.10 |
| Per Syndrome Mean: Frequent/Overall | 40.34 | 60.38 | 69.24 | 79.58 |
| Per Syndrome Mean: Rare/Overall | 18.86 | 30.36 | 36.03 | 47.50 |
| On AIDY test data (with AIDY gallery) |  |  |  |  |
| Category | Top 1 | Top 5 | Top 10 | Top 30 |
| Per Syndrome Mean: Frequent/Frequent | 43.38 | 67.82 | 76.58 | 86.23 |
| Per Syndrome Mean: Rare/Rare | 33.33 | 62.47 | 78.85 | 97.47 |
| Per Syndrome Mean: Frequent/Overall | 41.85 | 66.35 | 73.39 | 85.67 |
| Per Syndrome Mean: Rare/Overall | 20.07 | 41.24 | 51.4 | 70.89 |
| On AIDY test data with both GMDB and AIDY gallery |  |  |  |  |
| Category | Top 1 | Top 5 | Top 10 | Top 30 |
| Per Syndrome Mean: Frequent/Frequent | 44.00 | 69.53 | 78.44 | 89.69 |
| Per Syndrome Mean: Rare/Rare | 35.05 | 59.96 | 70.00 | 87.75 |
| Per Syndrome Mean: Frequent/Overall | 42.51 | 67.06 | 74.64 | 85.49 |
| Per Syndrome Mean: Rare/Overall | 22.74 | 37.79 | 43.14 | 64.03 |

**Table S5.**
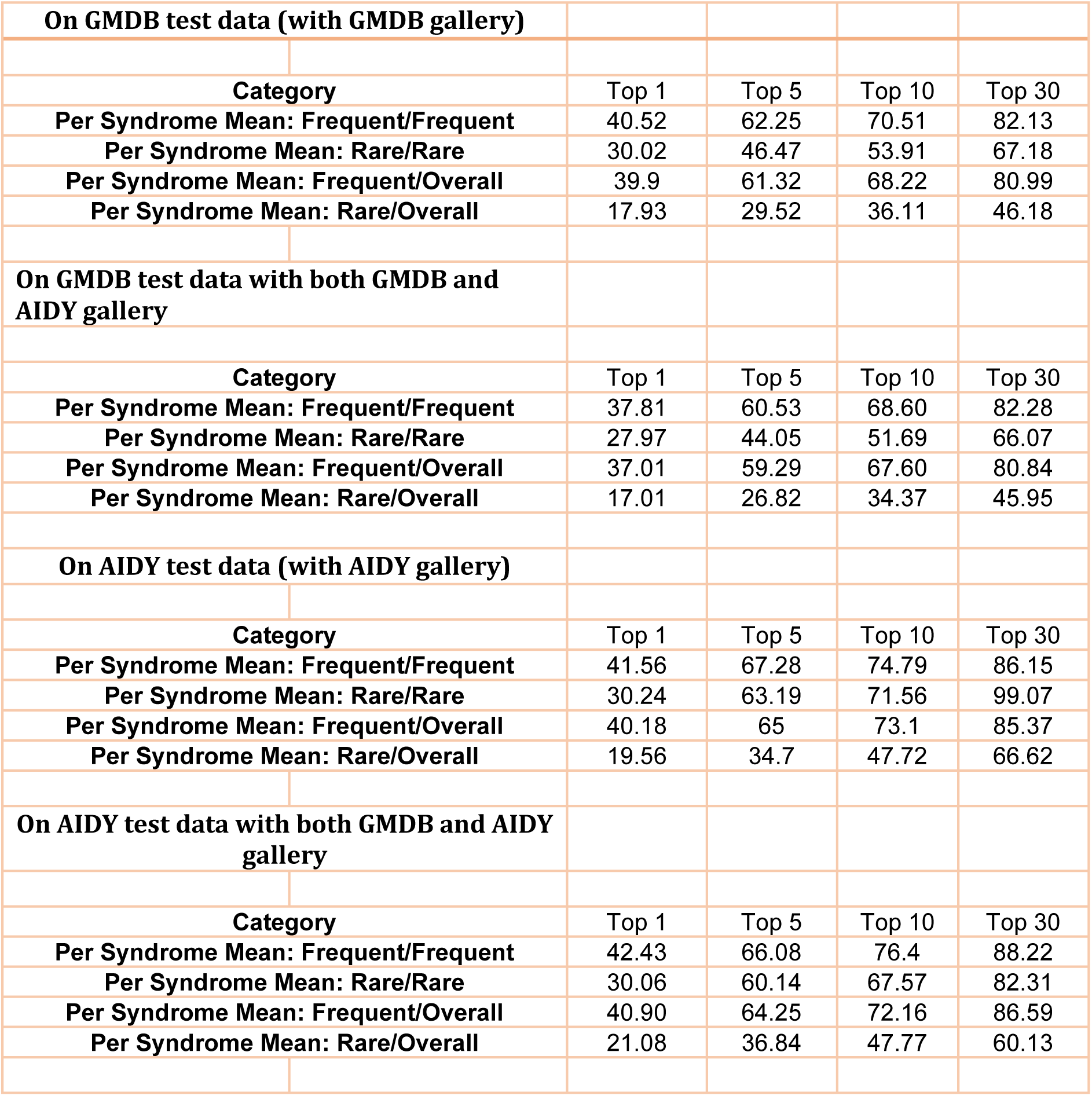
Performance of Swarm model.

|  |  |  |  |  |
| --- | --- | --- | --- | --- |
| **On GMDB test data (with GMDB gallery)** |  |  |  |  |
| **Category** | **Top 1** | **Top 5** | **Top 10** | **Top 30** |
| **Per Syndrome Mean: Frequent/Frequent** | 40.52 | 62.25 | 70.51 | 82.13 |
| **Per Syndrome Mean: Rare/Rare** | 30.02 | 46.47 | 53.91 | 67.18 |
| **Per Syndrome Mean: Frequent/Overall** | 39.9 | 61.32 | 68.22 | 80.99 |
| **Per Syndrome Mean: Rare/Overall** | 17.93 | 29.52 | 36.11 | 46.18 |
| **On GMDB test data with both GMDB and AIDY gallery** |  |  |  |  |
| **Category** | **Top 1** | **Top 5** | **Top 10** | **Top 30** |
| **Per Syndrome Mean: Frequent/Frequent** | 37.81 | 60.53 | 68.60 | 82.28 |
| **Per Syndrome Mean: Rare/Rare** | 27.97 | 44.05 | 51.69 | 66.07 |
| **Per Syndrome Mean: Frequent/Overall** | 37.01 | 59.29 | 67.60 | 80.84 |
| **Per Syndrome Mean: Rare/Overall** | 17.01 | 26.82 | 34.37 | 45.95 |
| **On AIDY test data (with AIDY gallery)** |  |  |  |  |
| **Category** | **Top 1** | **Top 5** | **Top 10** | **Top 30** |
| **Per Syndrome Mean: Frequent/Frequent** | 41.56 | 67.28 | 74.79 | 86.15 |
| **Per Syndrome Mean: Rare/Rare** | 30.24 | 63.19 | 71.56 | 99.07 |
| **Per Syndrome Mean: Frequent/Overall** | 40.18 | 65 | 73.1 | 85.37 |
| **Per Syndrome Mean: Rare/Overall** | 19.56 | 34.7 | 47.72 | 66.62 |
| **On AIDY test data with both GMDB and AIDY gallery** |  |  |  |  |
| **Category** | **Top 1** | **Top 5** | **Top 10** | **Top 30** |
| **Per Syndrome Mean: Frequent/Frequent** | 42.43 | 66.08 | 76.4 | 88.22 |
| **Per Syndrome Mean: Rare/Rare** | 30.06 | 60.14 | 67.57 | 82.31 |
| **Per Syndrome Mean: Frequent/Overall** | 40.90 | 64.25 | 72.16 | 86.59 |
| **Per Syndrome Mean: Rare/Overall** | 21.08 | 36.84 | 47.77 | 60.13 |

**Table S6.**
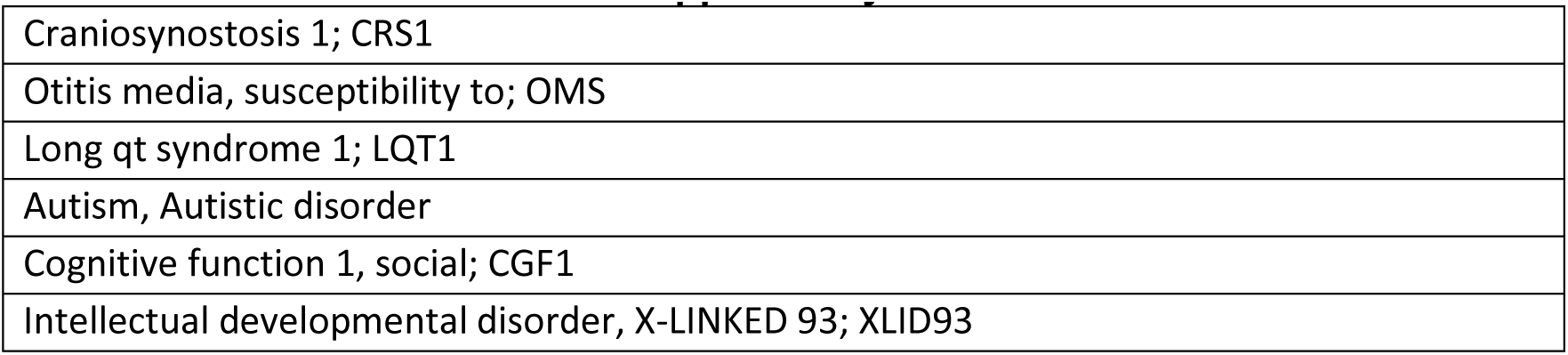
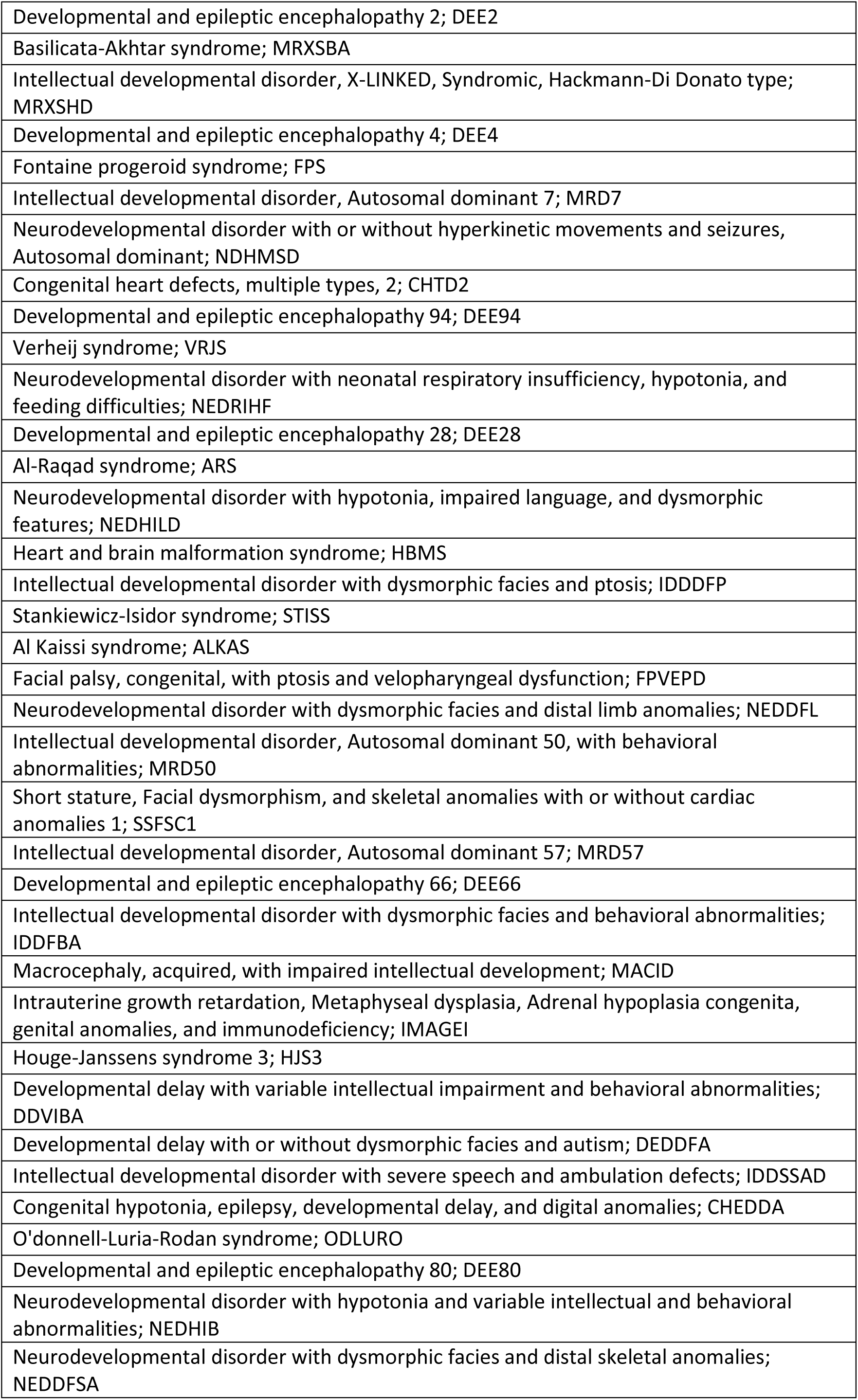
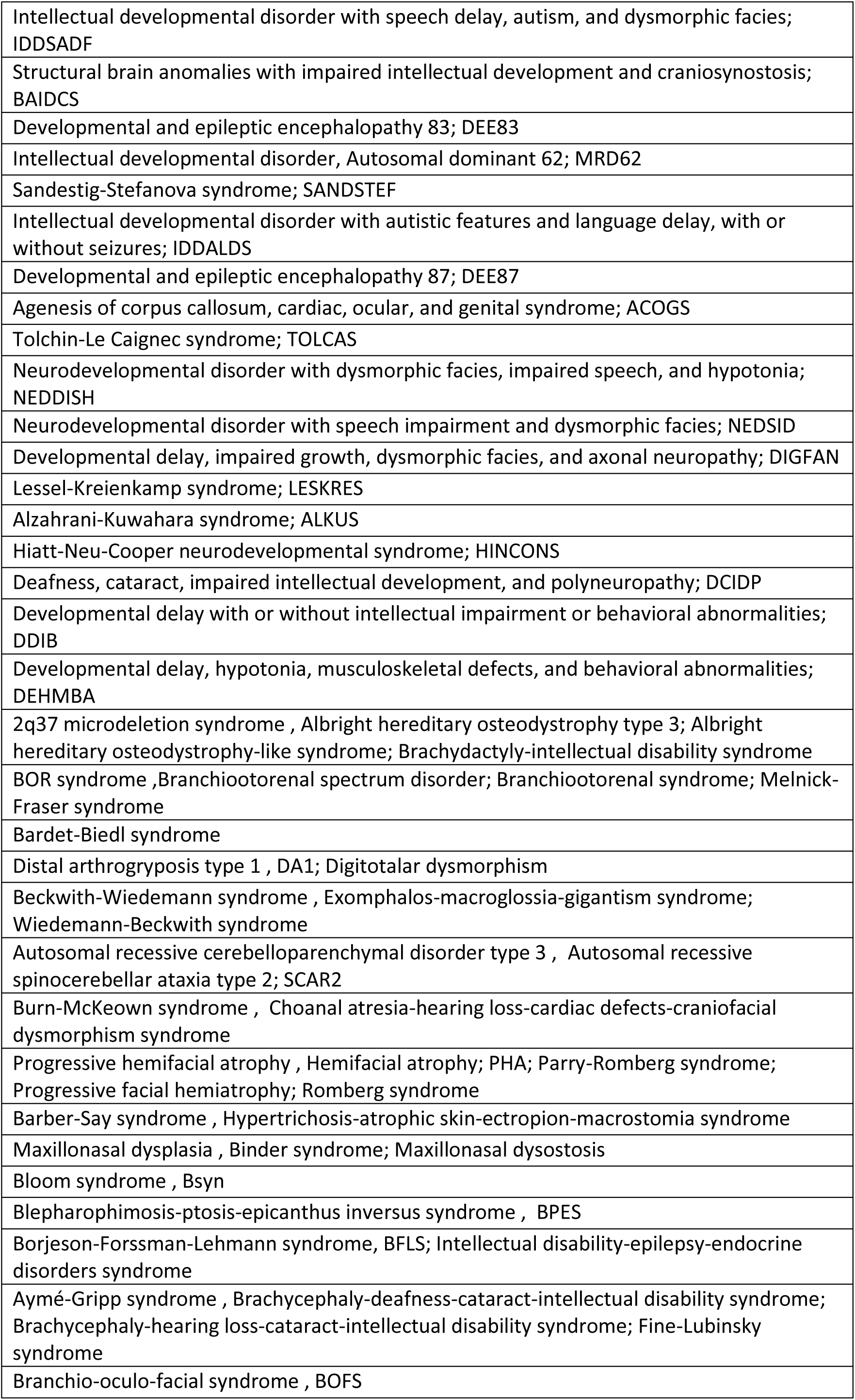
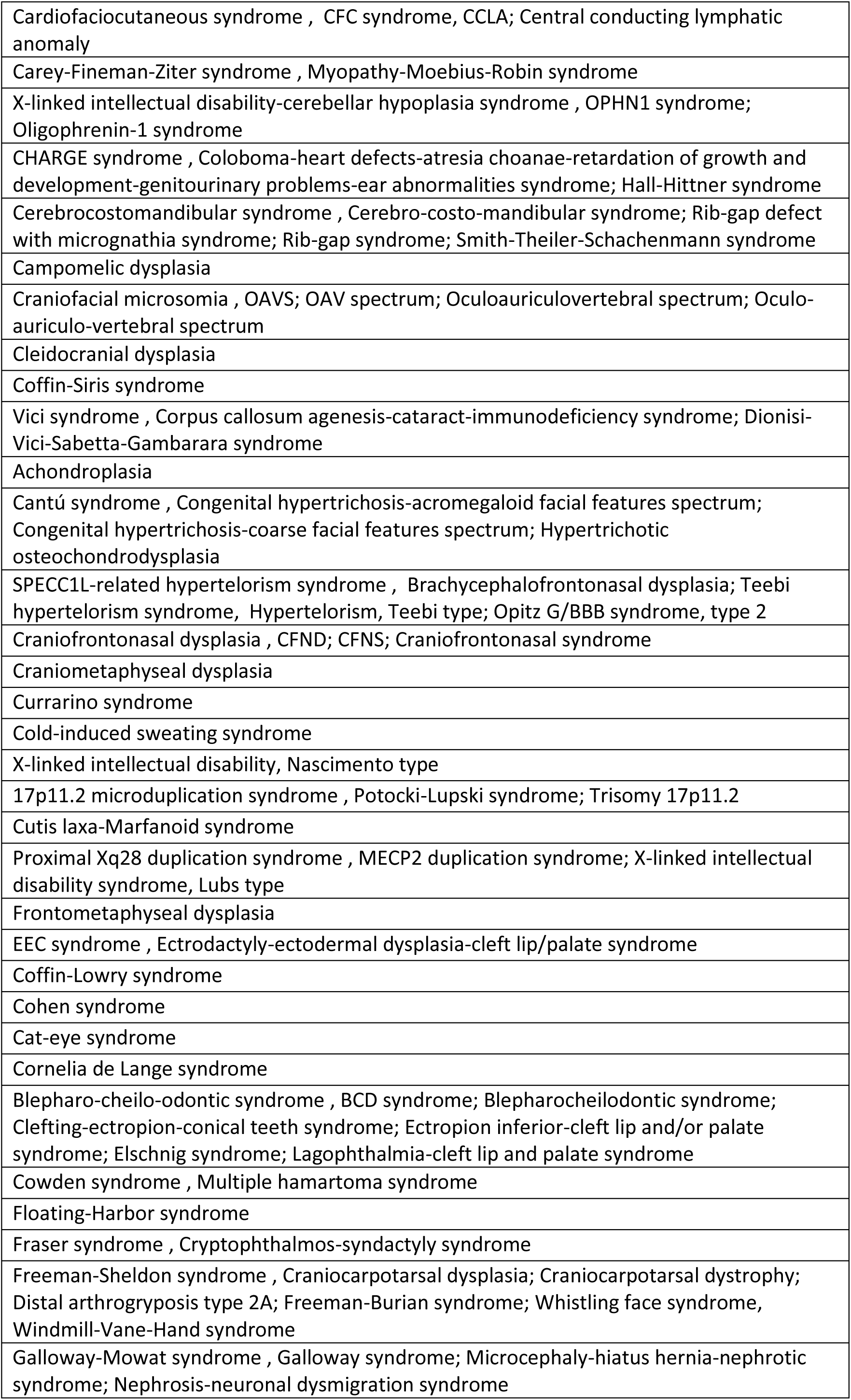
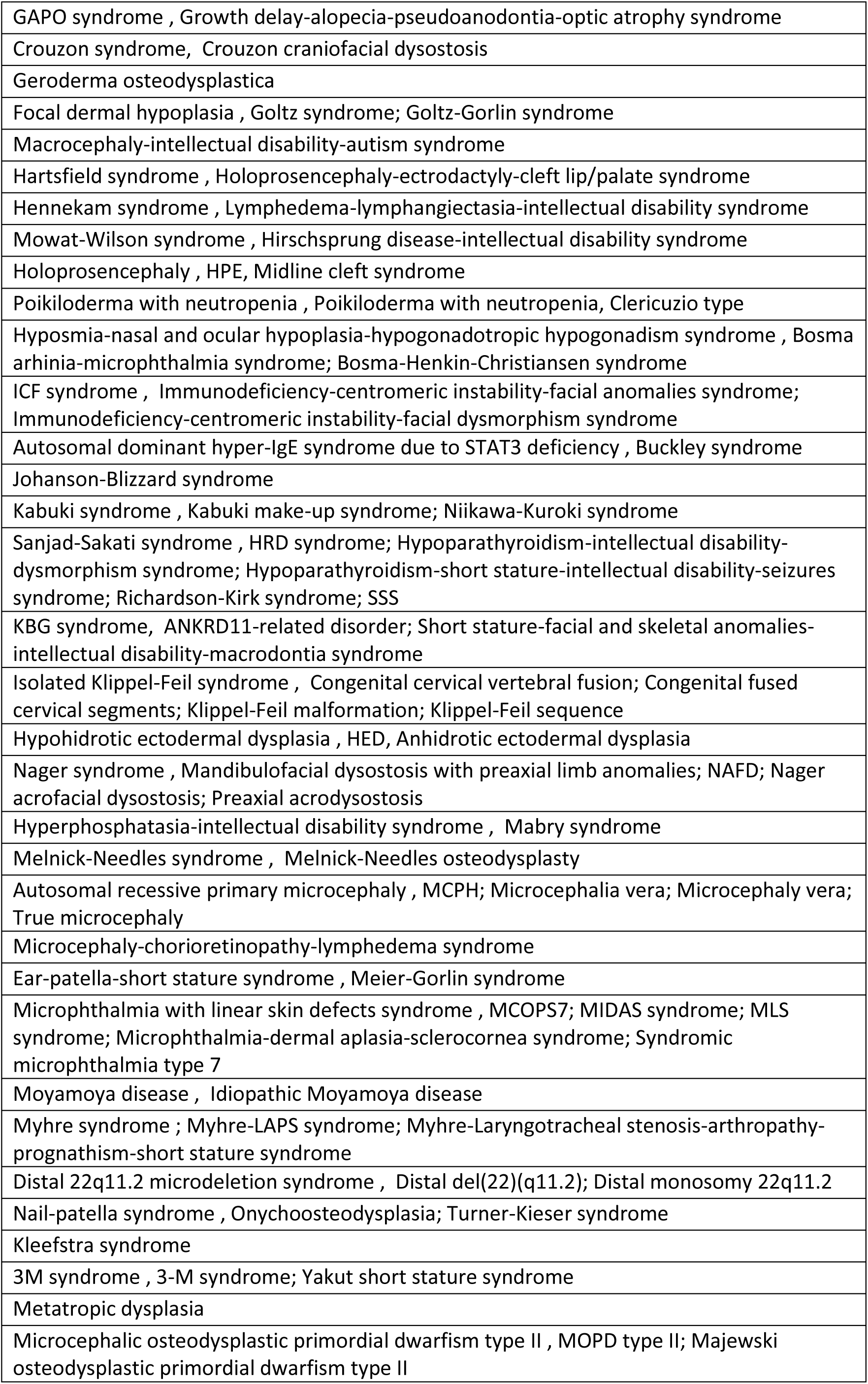
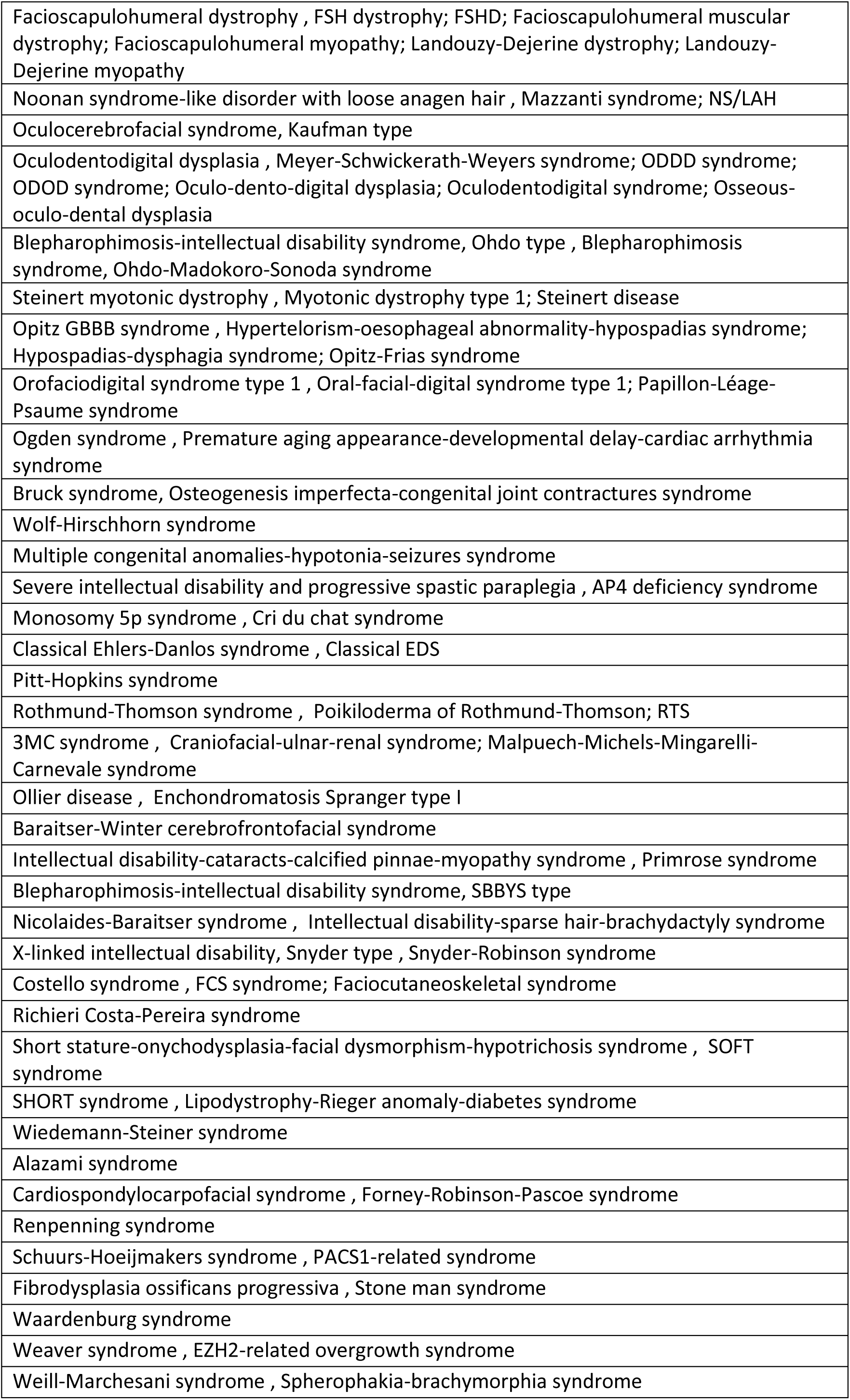
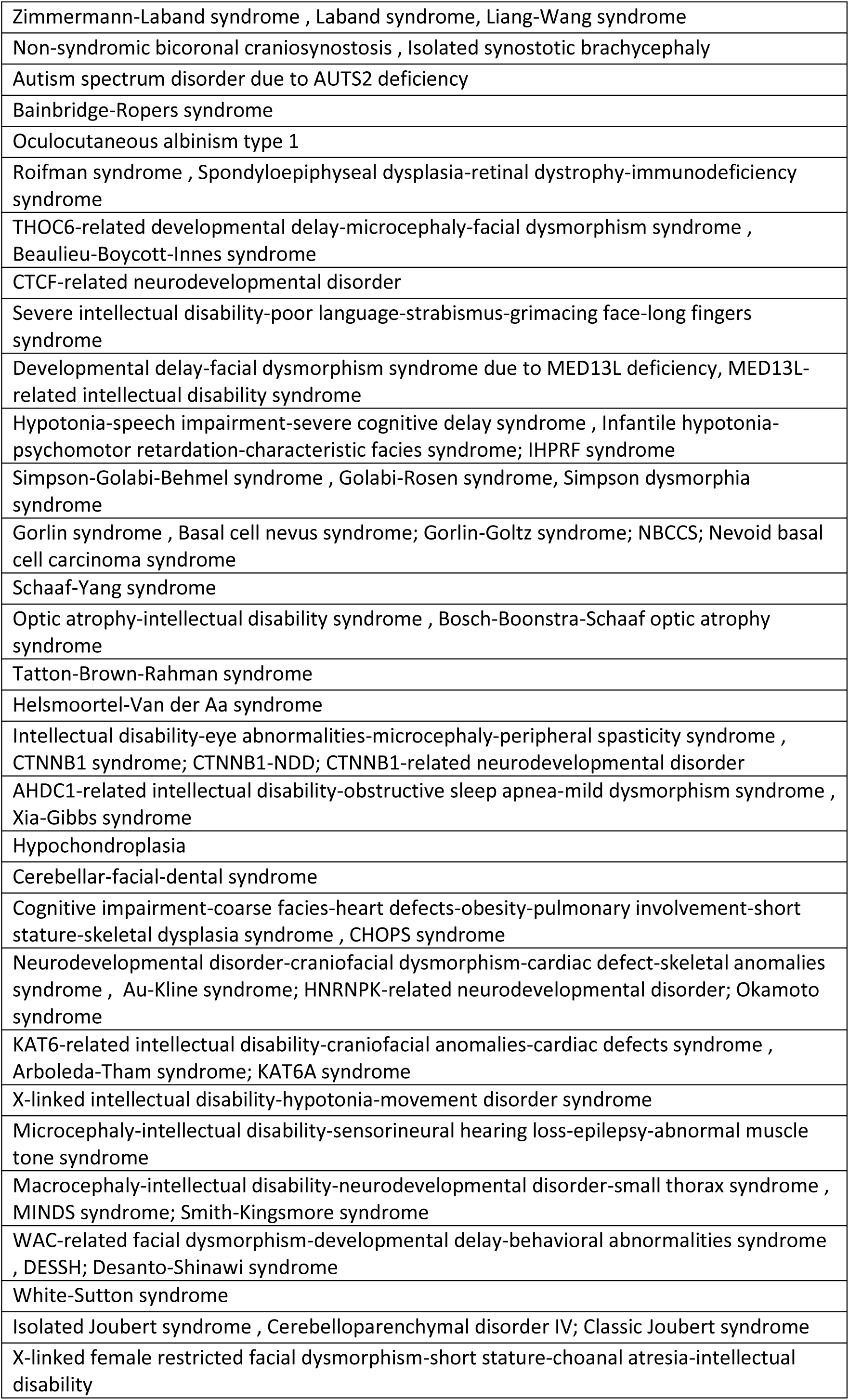
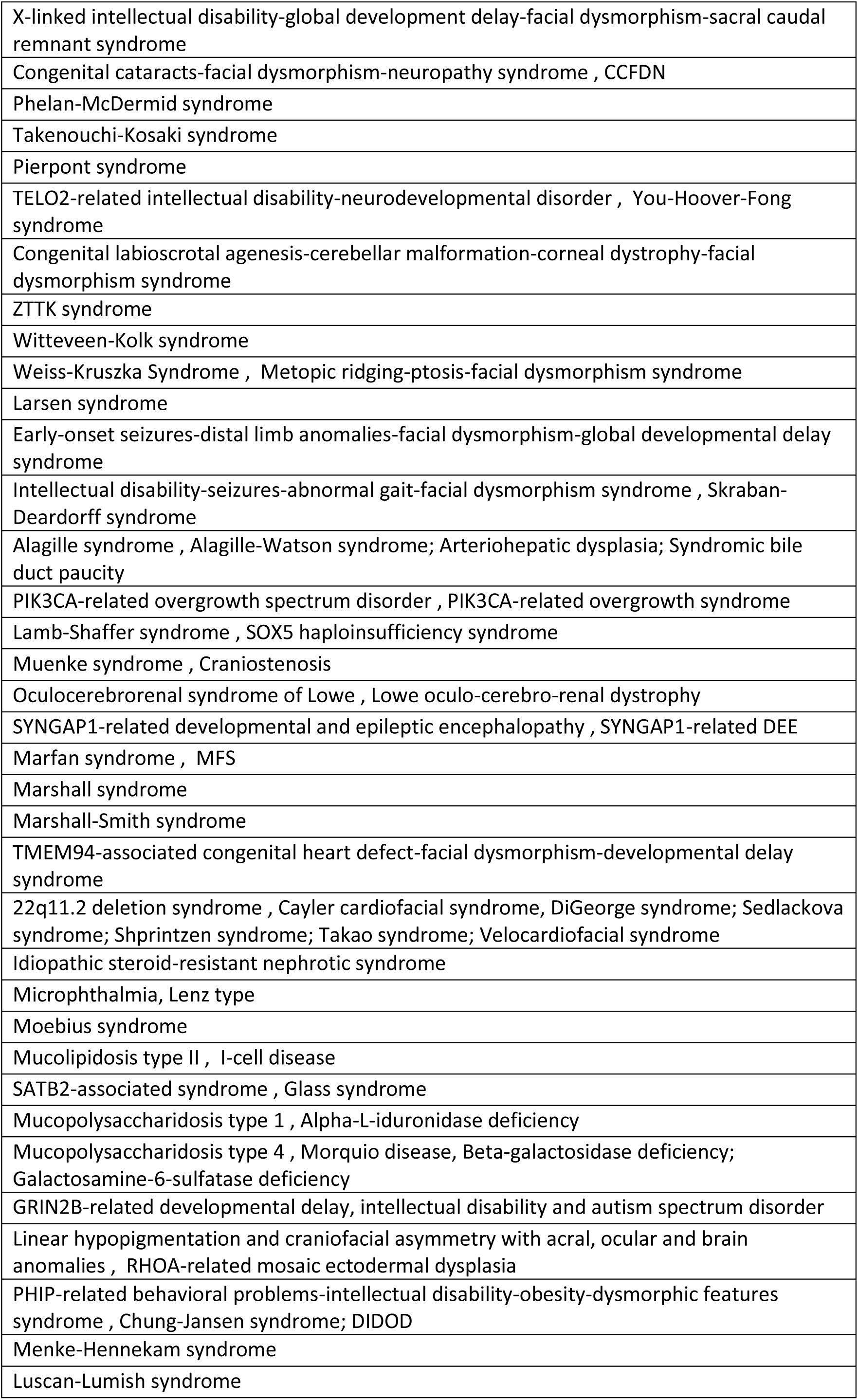
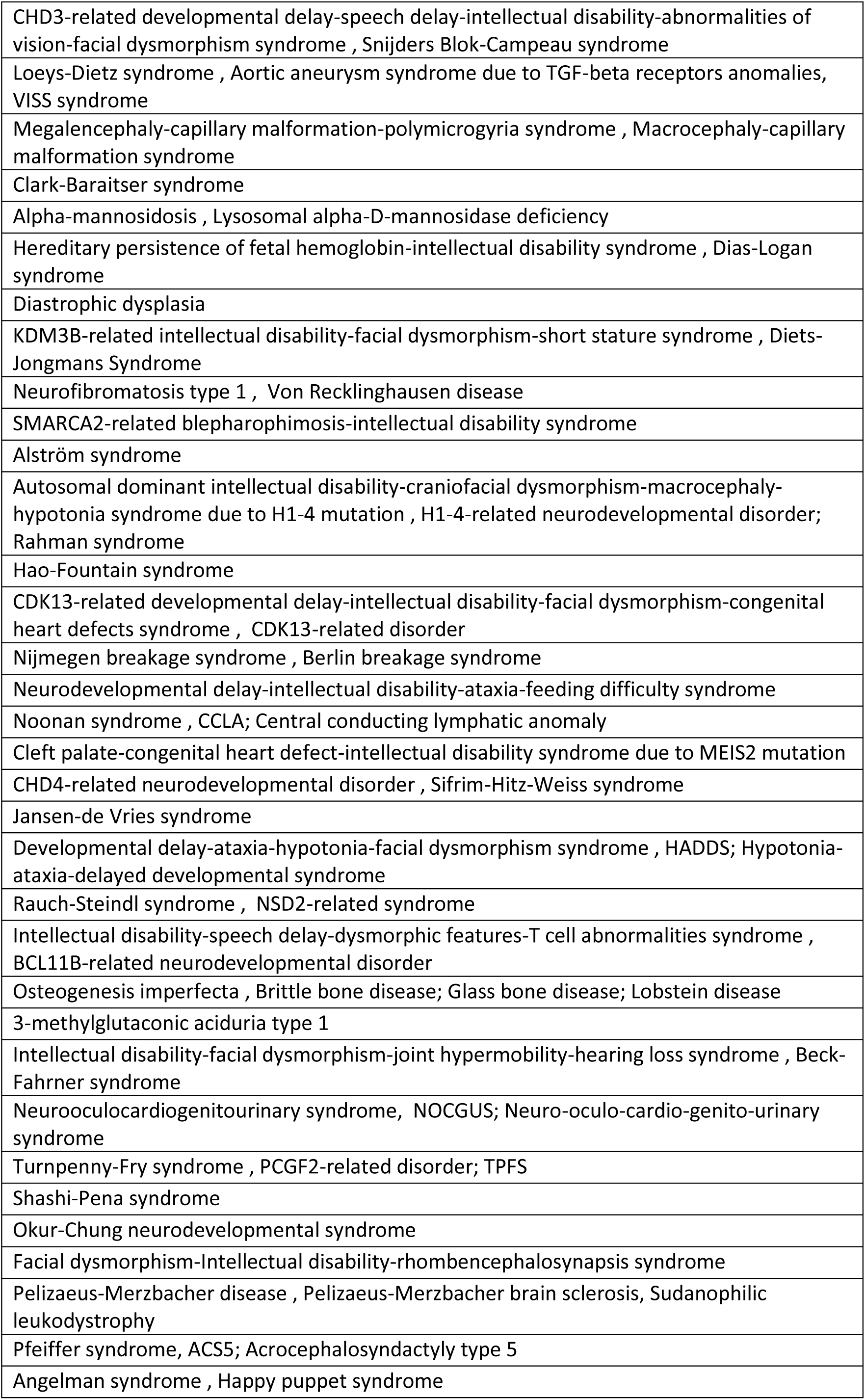
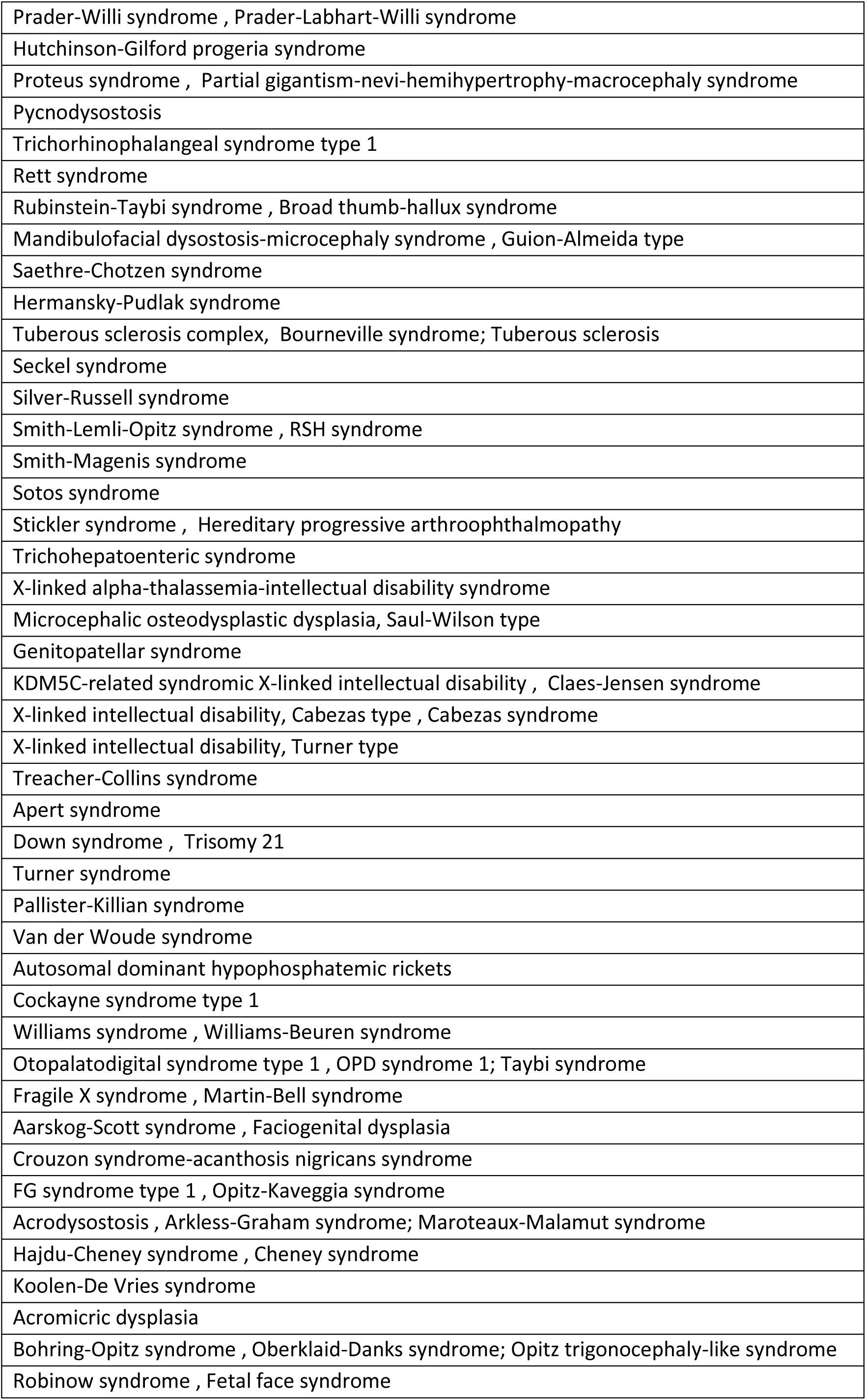
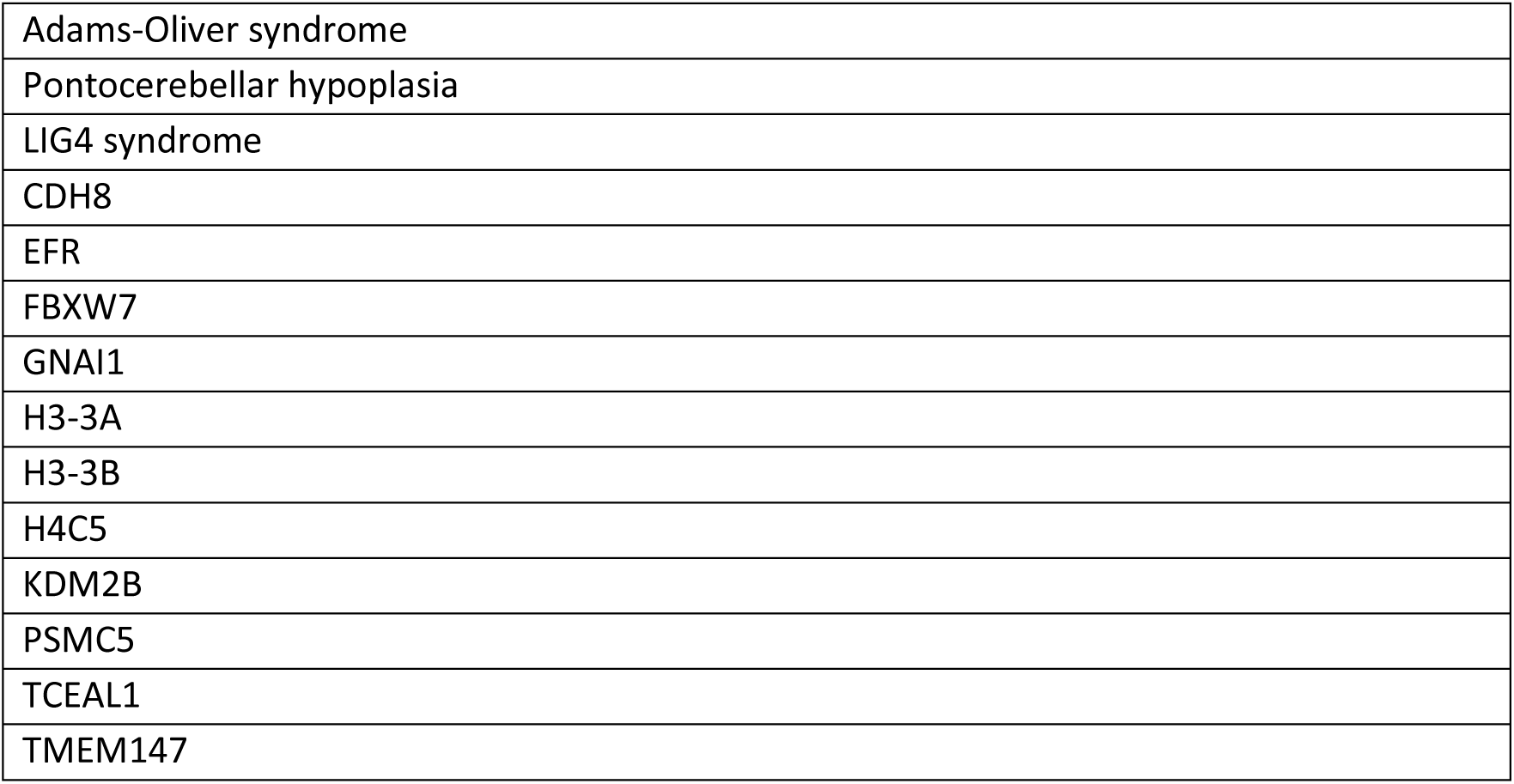
List of diseases supported by current Swarm model

